# A decade of shifting cholera burden in Africa and its implications for control: a statistical mapping analysis

**DOI:** 10.1101/2024.09.23.24314072

**Authors:** Javier Perez-Saez, Qulu Zheng, Joshua Kaminsky, Kaiyue Zou, Maya N. Demby, Christina Alam, Daniel Landau, Rachel DePencier, Jose Paulo M. Langa, Roma Chilengi, Placide Welo Okitayemba, Godfrey Bwire, Linda Esso, Armelle Viviane Ngomba, Nicole Fouda Mbarga, Emmanuel Wandera Okunga, Sebastian Yennan, Fred Kapaya, Stephen Ogirima Ohize, Adive Joseph Seriki, Sonia T. Hegde, Mustafa Sikder, Justin Lessler, Abhirup Datta, Andrew S. Azman, Elizabeth C. Lee

## Abstract

**Background:** The World Health Organization declared a global cholera emergency in 2023 due to an increase in cholera outbreaks, with most cholera-associated deaths reported in Africa. Characterizing large-scale burden patterns can help with monitoring progress in cholera control and targeting interventions.

**Methods:** We modeled the mean annual incidence of suspected cholera for 2011-2015 and 2016-2020 on a 20 km by 20 km grid across Africa using a global cholera database and spatial statistical models. We then examined how 2011-2020 incidence is associated with post-2020 cholera occurrence and investigated the potential reach of prospective interventions when prioritized by past incidence.

**Findings:** Across 43 African countries mean annual incidence rates remained steady at 11 cases per 100,000 population through both periods. Cholera incidence shifted from Western to Eastern Africa, and we estimated 125,701 cases annually (95% CrI: 124,737-126,717) in 2016-2020. There were 296 million (95% CrI: 282-312 million) people living in high-incidence second-level administrative (ADM2) units (≥ 10 cases per 100,000 per year) in 2020, of which 135 million experienced low incidence (<1 per 100,000) in 2011-2015. ADM2 units with sustained high incidence in Central and Eastern Africa from 2011-2020 were more likely to report cholera in 2022-2023, but cases were also reported in sustained low ADM2 units. Targeting the 100 million highest burden populations had potential to reach up to 63% of 2016-2020 mean annual cases but only 37% when targeting according to past 2011-2015 incidence.

**Interpretation:** By revealing the changing spatial epidemiology of cholera in Africa, these 10-year subnational estimates may be used to project OCV demand, characterize the potential of targeting interventions based on past burden, and track progress towards disease control goals.

## Introduction

Cholera has long been recognized as a major global public health issue, and it remains a significant cause of morbidity and mortality in low- and middle-income countries. The World Health Organization (WHO) declared a global cholera emergency in January 2023, prompted by an unexpected increase in cases in both recently cholera-free and acknowledged cholera-endemic areas ^1^ that reported over 800,000 cases and nearly 6,000 deaths between January 2023 and March 2024 ^2^. The emergency coincided with a prolonged shortage of oral cholera vaccine (OCV) which led WHO to suspend the recommended 2-dose course in favor of 1-dose reactive campaigns, despite evidence for lower sustained protection from the 1-dose regime ^3^. The WHO African region has witnessed over 4,500 deaths during the emergency period and consistently experiences high cholera burden even in non-emergency periods, with roughly 100,000 of 473,000 globally reported cases in 2022 ^2,4,5^.

The 2023 emergency underscores the challenge of realizing the vision for global cholera control to which WHO member states committed at the 71st World Health Assembly in 2018 ^6^. In this five-year interval, the Global Task Force on Cholera Control (GTFCC) and governments have made significant progress in cross-cutting coordination, developing and implementing guidance for the management and surveillance of cholera and its associated risk factors, and expanding the use of OCVs ^7–14^. Nevertheless, additional effort is required to generate sustained progress towards global cholera control, some of which could be achieved through efficient regional targeting of control measures like improvements in water and sanitation infrastructure, case-area targeted interventions (CATIs), and mass OCV campaigns ^15–19^. Current GTFCC recommendations on targeting interventions rely on past incidence, and significant changes in spatial burden patterns may compromise their efficiency ^20^. As such, estimating the burden of disease across large geographic scales and how they change over time can be of key importance, both for tracking advancement towards broad disease control objectives ^21–23^ and identifying priority areas for regional planning and coordination. African countries have led the charge in developing multi-year cholera control plans ^10–13^, and several countries are working to update plans that are now several years old.

Leveraging a global database of cholera surveillance data with spatial statistical models, our study presents 20 km by 20 km maps of estimated medically-attended suspected cholera incidence in Africa from 2011-2015 and 2016-2020. Over this 10-year period, we examine changes in the overall burden, spatial distribution, and number of people living in high incidence areas over time. In addition, we model the association between 2011-2020 cholera incidence and the spatial distribution of cholera in the post-2020 period. Finally we assess the potential reach of targeting intervention when prioritizing by past cholera incidence.

## Methods

### Cholera incidence data

We curated a global cholera incidence database of national and subnational surveillance data and other reports from Ministries of Health (MOHs), GTFCC partners, and other public data sources for countries in Africa from 2011 through 2020, which included a comprehensive online search for national and subnational cholera outbreak reports for every modeled country and year. Shapefiles were obtained from MOHs, WHO country or regional offices, unified or curated sources like GADM, geoboundaries, and Grid3, and other online sources ^24–26^, and linked to observation locations. Suspected cholera case definitions varied by data source and were oftentimes not stated, but were commonly variations of the recommended WHO suspected case definition, such as “any patient presenting with or dying from acute watery diarrhea” and “a patient aged 2 years or more develops acute watery diarrhea with or without vomiting.” (cf. Table S1 for a complete list).

All countries in Africa that had at least one national-level report of suspected cholera (including zero) in both periods of analysis were modeled (Table S2). We developed a processing pipeline to format and harmonize raw data inputs (which covered a wide range of temporal and spatial scales) for our statistical mapping modeling framework (Figures S1-S2, *SM - Cholera data sources and data processing*).

### Other spatial data sources

Gridded annual 1 km x 1 km population estimates were taken from the unconstrained global mosaic WorldPop population counts dataset^27^. The gridded estimates were then scaled such that the total country population matched the respective annual estimate from the 2022 revision of the UN World Population Prospects ^28^. For map visualizations, we accessed major water bodies in Africa published by the Regional Centre for Mapping of Resources for Development and AQUASTAT programme of the Food and Agriculture Organization of the United Nations ^29,30^.

### Statistical burden mapping model

We developed a hierarchical Bayesian modeling framework that accounts for spatio-temporal heterogeneity in underlying suspected cholera incidence and variability and overlap in the spatial and temporal scales of case reports within and across data sources.

We modeled each country and time period (2011-2015 and 2016-2020) separately. The process model for most country-periods consisted of a log-linear model of annual cholera incidence rates over a 20 km by 20 km grid accounting for spatial auto-correlation and inter-annual variability. Spatial auto-correlation was implemented through a directed acyclic graph auto-regressive (DAGAR) prior for the spatial random effects which has improved performance relative to traditional spatial priors in disease mapping ^31^. Temporal variability was modeled through annual temporal random effects. Yearly observations and temporally censored observations contributed to separate parts of the likelihood. The process models for country-periods with no or minimal subnational data were modified to improve interpretability of results and model performance (cf. Table S3 for model settings by country and *SM - Incidence modeling framework* for model details). The modeled mean incidence rate for an area corresponding to a reported case count was derived as the weighted average of the incidence rates of grid cells covering that area. Observations overlapping in space and time were treated as independent measurements of the same underlying incidence rate. To account for reporting variability across data sources, we used a negative-binomial observation model for the case counts with inferred administrative level-specific overdispersion parameters (*SM - Incidence modeling framework*).

Posterior samples were drawn with Hamiltonian Monte Carlo (HMC) as implemented in the Stan programming language ^32^. Sampler convergence was assessed visually through the inspection of trace plots and observation-level Rhat statistics ^33^. Model fit was evaluated through scatterplots of true and fitted observations and posterior retrodictive checks of the posterior coverage of observations by administrative level ^34^ (Figures S3-S12). Gridded outputs were post-processed for mean annual incidence, mean annual incidence rate, incidence rate ratio, population in ADM2 units in 5-year and 10-year incidence categories, assignment of ADM2 units in 5-year and 10-year incidence categories at the administrative unit levels 0 and 2 (ADM0 and ADM2, sometimes called country and district-level, respectively), region, and continent scales for analysis (*SM - Incidence modeling post-processing*).

### Definition of 5-year and 10-year incidence categories

We identified six categories pertaining to the 5-year mean annual incidence: <1, [1, 10), [10, 20), [20, 50), [50, 100), and ≥100 cholera cases per 100,000 people per year. For a given posterior sample, ADM2 units were assigned to the most severe 5-year incidence category where at least 10% of the unit’s population or 100,000 people (in 2020-adjusted population sizes) were living, according to the modeled 20 km by 20 km grid cell estimates (cf. *SM - Incidence modeling post-processing* for details). This incidence category assignment procedure was designed to identify locations that may make high impact targets for public health intervention ^4^. We used the labels “very high incidence” to refer to the category of ≥100 cases per 100,000 population per year, “high incidence” for ≥10 per 100,000, and “low incidence” for <1 per 100,000.

We also used a 10-year incidence categorization at the ADM2 level that combines the incidence categories in each 5-year period (2011-2015 and 2016-2020). We defined four 10-year incidence categories: “sustained high” for ADM2 units classified as high incidence (≥10 cases per 100,000 people per year) in both periods, “history of high” for ADM2 units classified as high incidence in at least one period, “sustained low” for ADM2 units classified as <1 case per 100,000 per year in both periods, and “history of moderate” for all other combinations.

### Statistical analysis of 2022-2023 cholera occurrence

We evaluated the association between the 10-year incidence categories (from 2011 to 2020) and cholera occurrence as reported in 2022-2023 WHO external cholera situation reports 1 through 10 ^1,35–43^, complemented with country-specific situation reports for areas not displayed in WHO reports ^44–49^ (Table S5). The extracted data corresponded primarily to cholera reported between January 2022 and December 2023, with limited additional data in surrounding months due to the temporal reporting resolution. For this analysis, locations were determined to have cholera if one or more suspected cholera cases were reported in any of the above-described situation reports. We then spatially joined extracted locations to the set of ADM2 units used for summarizing the cholera incidence mapping results (*SM - Analysis of 2022-2023 cholera occurrence*).

As the situation reports provided limited subnational case data, we evaluated the association between 10-year incidence categories and recent cholera occurrence in a Bayesian modeling framework. The model consisted of a hierarchical logistic regression that accounted for the probability of failing to detect cholera (false negatives), reporting of cases at multiple administrative levels, as well as partial pooling of country-specific parameters values at the regional and continental levels (*SM - Analysis of 2022-2023 cholera occurrence*). Inference was performed with HMC as implemented in the Stan programming language ^32^.

### Assessing intervention reach when prioritizing targets by cholera incidence

We assessed the potential reach of interventions when targeted based on cholera incidence through two analyses; both analyses prioritized ADM2 units by decreasing incidence category and decreasing population size within incidence categories and examined what proportion of 1) mean annual cholera cases or 2) population living in ADM2 units with 2022-2023 cholera occurrence would have been reached under different targeting strategies (See *SM - Assessing intervention reach when prioritizing targets by cholera incidence* for details). “Prospective” targeting used past incidence categories to target future interventions, while “oracle” targeting prioritized interventions based on incidence categories from the same period.

### Inclusion and ethics statement

The Institutional Review Board (IRB) at Johns Hopkins Bloomberg School of Public Health (BSPH) determined that secondary analysis of data from the global cholera incidence database was exempt (BSPH IRB No. 27682) and no other institutional approvals were sought.

Twelve authors (J.P.M.L, R.C., P.W.O., G.B., L.E., A.V.N., N.F.M., E.W.O., S.Y., F.K., S.O.O., A.J.S.) contribute to public health activities in cholera-affected low- and middle-income countries (LMICs). They provided feedback on the interpretation and application of this work based on their expertise in cholera surveillance and control in LMIC contexts. We fully endorse the Nature Portfolio journals’ guidance on LMIC authorship and inclusion, and are strongly committed to the inclusion of more researchers and decision-makers from LMICs in future related work.

### Data and code availability

Publicly available cholera incidence datasets may be viewed and accessed from cholera-taxonomy.middle-distance.com. Metadata for confidential datasets is available upon request. Data processing and modeling code is publicly available on Github at https://github.com/HopkinsIDD/cholera-mapping-pipeline, release v1.0.

## Results

Our analysis dataset consisted of 30,211 distinct reports of suspected cholera cases (henceforth “observations”) between 2011 and 2020 from 807 distinct data sources. Observations covered 4,574 unique geographical areas (henceforth “locations”) and spanned seven administrative levels (863 at national and 29,239 at subnational levels) across 43 countries in Africa with cholera reporting for at least one year (Table S2). Both time periods (2011-2015 and 2016-2020) had similar numbers of observations, but the number of unique locations and data sources was larger in the more recent period (Table S2, Figures S13-S14). Subnational observation coverage varied considerably between countries, with the area covered by ADM2 or lower observations in at least one year ranging from 0.03% (Namibia) to 100% (11 countries) with a median of 85% (Figure S15). After temporal aggregation to the yearly scale, 73% of observations (22,080 of 30,211) covered at least 8 months of the year and were considered full-year observations in our modeling framework (*SM - Cholera data sources and data processing*, Table S6).

We estimated an annual average of 125,701 (95% Credible Interval, CrI: 124,737-126,717) suspected cholera cases across Africa in 2016-2020, which was an increase relative to 2011-2015 (105,753; 95% CrI: 104,550-107,005) (Fig 1A). At the regional level, the 2016-2020 case burden concentrated in Eastern and Central Africa, while the 2011-2015 case burden was more evenly distributed. Overall, 491 million out of 1.1 billion people in 2016-2020 (355 out of 960 million in 2011-2015) lived in 20 km x 20 km grid cells having more than 1 reported case per year, but these were spatially confined to only 17% of modeled grid cells in 2016-2020 (14% in 2011-2015) (Fig 1B, Figure S16).

**Figure 1.**
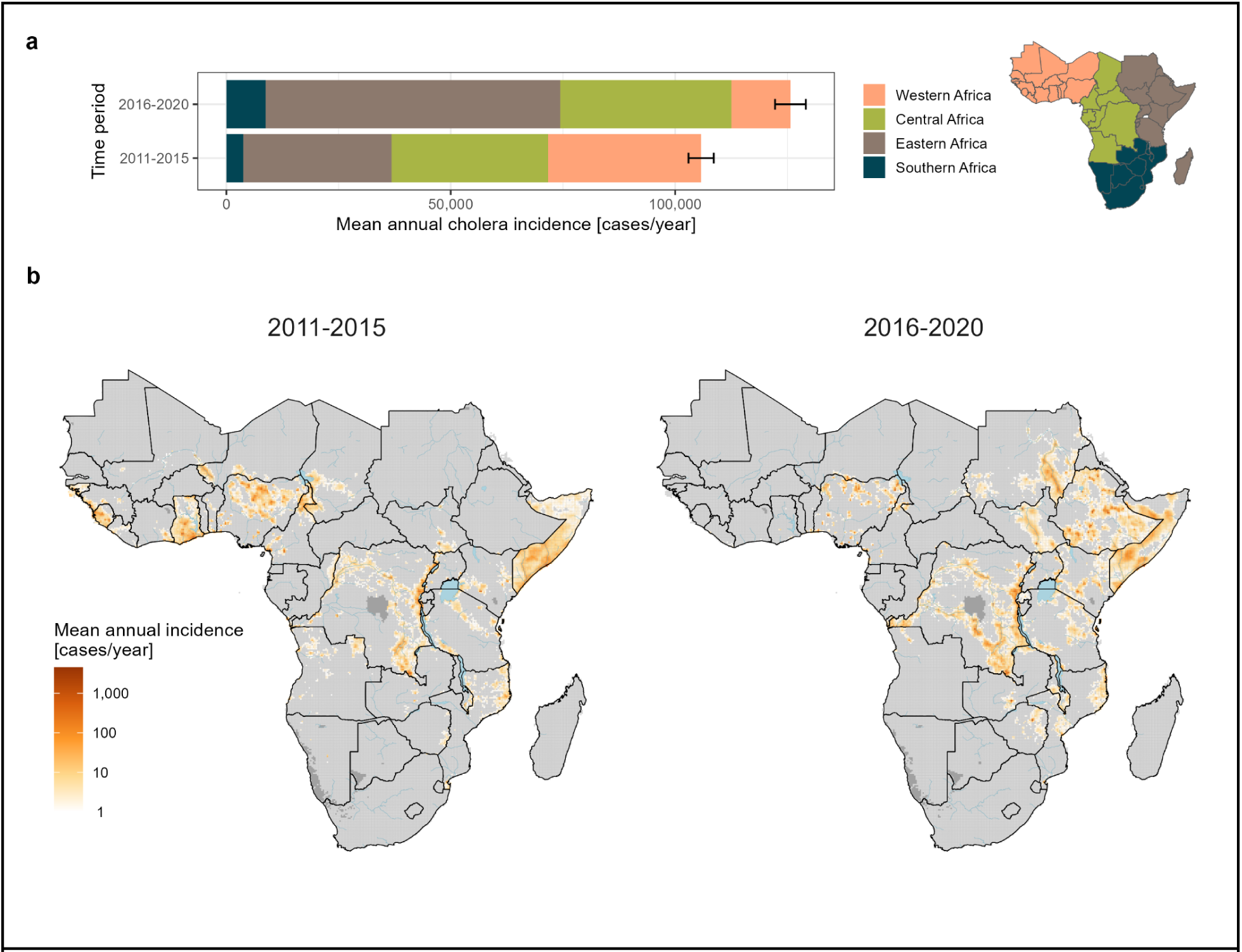
Mean annual suspected cholera incidence (cases per year) in Africa from 2011-2020. a) Mean annual suspected cholera incidence colored by region in Africa for two time periods (2011-2015 and 2016-2020). Error bars represent the 95% CrI for the posterior predictive samples of the continent-wide mean annual incidence. The colored map on the right is a legend depicting regions within Africa. b) Gridded 20 km by 20 km estimates of mean annual suspected cholera incidence in Africa from 2011-2015 (left) and 2016-2020 (right). Grid cells in light gray had a mean of less than 1 case per year, while those in dark gray were unmodeled due to zero population in the underlying population grid. Blue shaded areas represent major lakes and rivers in Africa.

The continent-wide mean annual incidence rate remained steady across both periods, hovering just above 11 cases per 100,000 population (Fig 2A). However, there were significant regional differences with increases in cholera burden in Eastern (incidence rate ratio, IRR 1.73, 95% CrI: 1.67-1.79) and Southern Africa (IRR 2.08, 95% CrI: 1.98-2.17) and a decrease in Western Africa (IRR 0.34, 95% CrI: 0.33-0.35). Overall, 13 countries had significant increases in burden (IRR>1) between the two periods and 24 had significant decreases (IRR<1) (Fig 2A). We observed subnational shifts in the spatial distribution of burden as well, including a total of 16 countries with both increases and decreases in their ADM2 incidence rates (Fig 2B).

**Figure 2.**
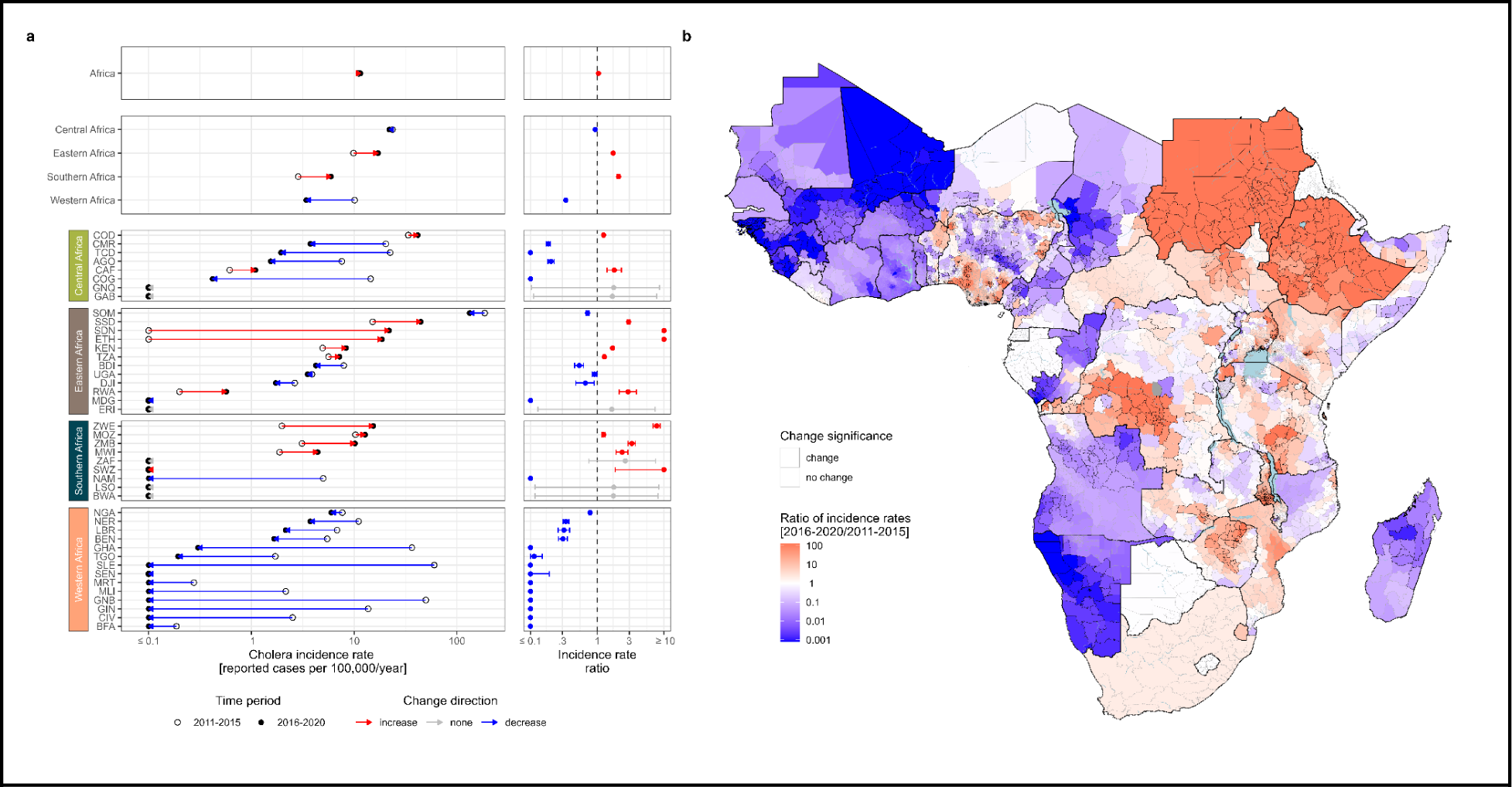
Changes in mean annual suspected cholera incidence rate (cases per population per year) in Africa from 2011-2020. a) The dot plots (left column) display changes in the posterior mean of the mean annual incidence rate from 2011-2015 (open circle) to 2016-2020 (filled circle) at the continent-level (top panel), region-level (second panel), and country-level listed by ISO3 code (four bottom panels). Arrows indicate the direction of change from 2011-2015 to 2016-2020, with red indicating increases and blue indicating decreases. Countries are ordered by decreasing 2016-2020 mean annual incidence rate within region-level panels. All changes shown in the figure were statistically significant except those in Botswana, Eritrea, Gabon, Equatorial Guinea, Lesotho, and South Africa. The incidence rate ratio plots (right column) display the mean (point) and 95% CrI (bars) for the posterior distribution of the mean annual incidence rates in 2016-2020 relative to 2011-2015. b) Posterior mean ratio of the mean annual incidence rates in 2016-2020 relative to 2011-2015 by ADM2 units. Units filled with red had higher mean rates and areas filled with blue had lower mean rates in the 2016-2020 period. ADM2 units outlined in dark gray had 95% CrIs completely above or below 1, respectively. ADM2 units outlined in light gray represent statistically non-significant differences. The following countries were not eligible to have statistically significant ADM2 changes due to limited subnational data in at least one of the two periods: Côte d’Ivoire, Djibouti, Ghana, Liberia, Rwanda, Senegal, Eswatini, South Africa. Areas in light blue represent large water bodies. All areas displayed were modeled.

In 2016-2020, we estimated there were 296 million (95% CrI: 282-312 million) people living in high incidence areas (≥10 per 100,000 population), among which 82 million (95% CrI: 72-91 million) experienced very high incidence (≥100 per 100,000 population) (Fig 3A). Most people in high incidence areas were located either in Eastern Africa (166 million, 95% CrI: 155-177 million) or Central Africa (66 million, 95% CrI: 60-72 million) (Fig 3A). We found that 764 of 4193 (18%) ADM2 units were assigned to high incidence categories in 2016-2020, and that these are concentrated in only 20 of 43 modeled countries (Fig 3B). Results for 2011-2015 are reported in the supplement (Figure S17-S18).

**Figure 3.**
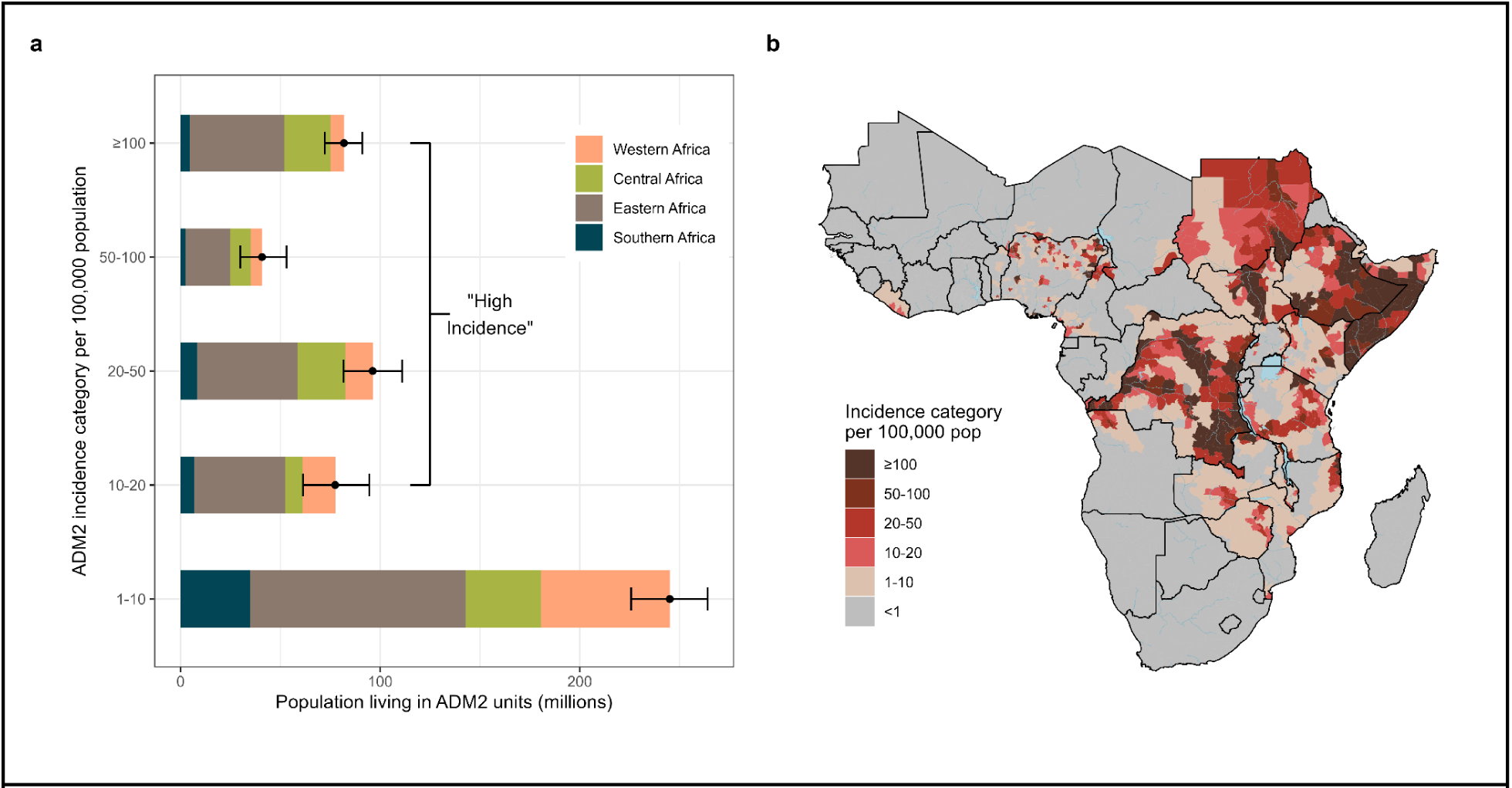
Population living in areas according to incidence category in 2016-2020. a) Mean (black point) and 95% credible interval (black bars) for district populations living in a given incidence category (per 100,000 population) across Africa (cf. Methods for incidence category definitions). Regional population contributions are indicated by fill colors and categories ≥10 per 100,000 population are labeled as “high incidence” categories. b) Continent-wide map showing assignment of incidence categories to ADM2 units by color. ADM2 units were assigned to an incidence category if 50% of posterior draws classified the ADM2 unit to the assigned color of incidence category or above. ADM2 units in gray had an incidence category of <1 per 100,000 population. Only modeled countries are displayed in the map.

Across the 2011-2020 period, we found that 105 million out of 1.1 billion people in Africa (10%) lived in 346 ADM2 units categorized as “sustained high” incidence (≥10 per 100,000 in both periods) (Fig 4A), across 17 countries located mostly in Eastern and Central Africa (Fig 4B). There were another 313 million (29%) in “history of high” incidence (≥10 per 100,000 in exactly one period). This included people living in ADM2 units with large swings in incidence between the two periods, notably 135 million experiencing low incidence in 2011-2015 and high incidence in 2016-2020 (versus 80 million in ADM2 units shifting from high to low incidence) (Fig 4A). Ten countries across Central, Eastern, and Western Africa had over 50% of their population living in areas with “sustained high” and “history of high” incidence (Fig 4B, Figure S19). Overall, only 342 million people (32%) lived in “sustained low” incidence ADM2 units (<1 per 100,000 in both periods) (Fig 4A).

**Figure 4.**
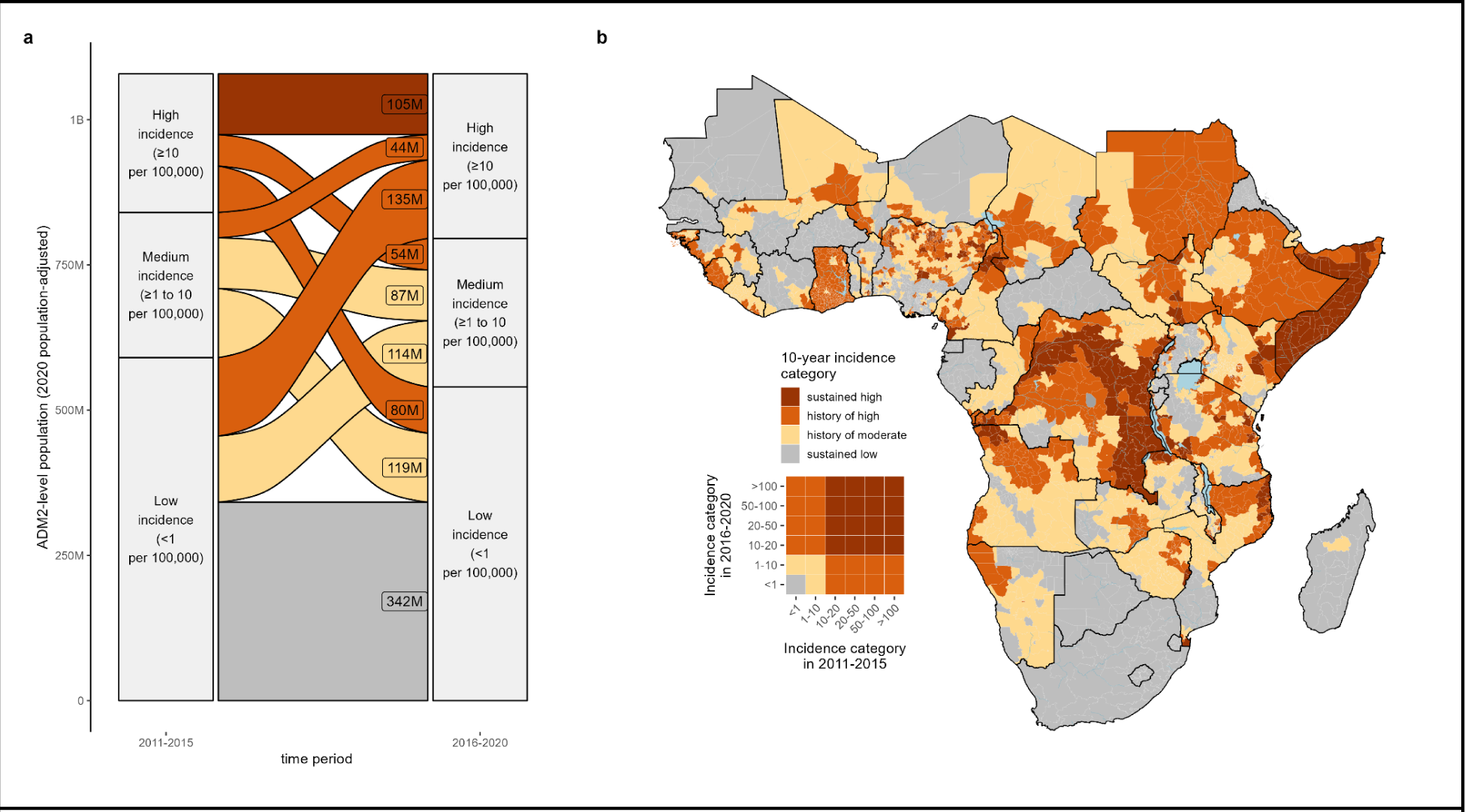
Ten-year incidence category of cholera burden in Africa across 2011-2020. a) Alluvial plot depicting changes in the number of people living in ADM2 units according to their 5-year incidence categories in 2011-2015 (left) and 2016-2020 (right), adjusted for 2020 population size. The flow colors indicate the 10-year (2011-2020) incidence category. The flow-specific labels indicate the number of people in ADM2 units according to their 2020 population size. b) Continent-wide map showing assignment of 10-year incidence categories to ADM2 units by color (cf. Methods for definition of 10-year incidence categories). The inset is a visual legend that translates the cross of two 5-year incidence categories into the 10-year incidence category. Grey represents ADM2 units with sustained low incidence. Only modeled countries are displayed in the map.

In 2022-2023, suspected cholera was reported in 502 geographic areas across 19 countries, among which 65.9% were at ADM2 level or lower (283 locations) (Fig 5A). The majority of locations with reported cholera were in “sustained high” and “history of high” ADM2 units (182 of 283 ADM2 units), although 35 “sustained low” ADM2 units also saw cholera cases (Fig 5B). Using statistical models that account for possible under-reporting, we found that the odds of 2022-2023 cholera occurrence tended to increase with the severity of the 10-year incidence category, although results were not statistically significant in all regions. The largest odds ratios were observed in the “sustained high” category in Central (median OR 75.4, 95% CrI: 2.7-3,875.0) and Eastern Africa (median OR 50.7, 95% CrI: 4.5-3,216.8) (Fig 5C). Cholera occurrence was predicted to be unlikely in “sustained low” ADM2 units except in Southern Africa (0.19, 95% CrI: 0.01-0.76). Country-specific odds ratios were also calculated (Figure S20).

**Figure 5:**
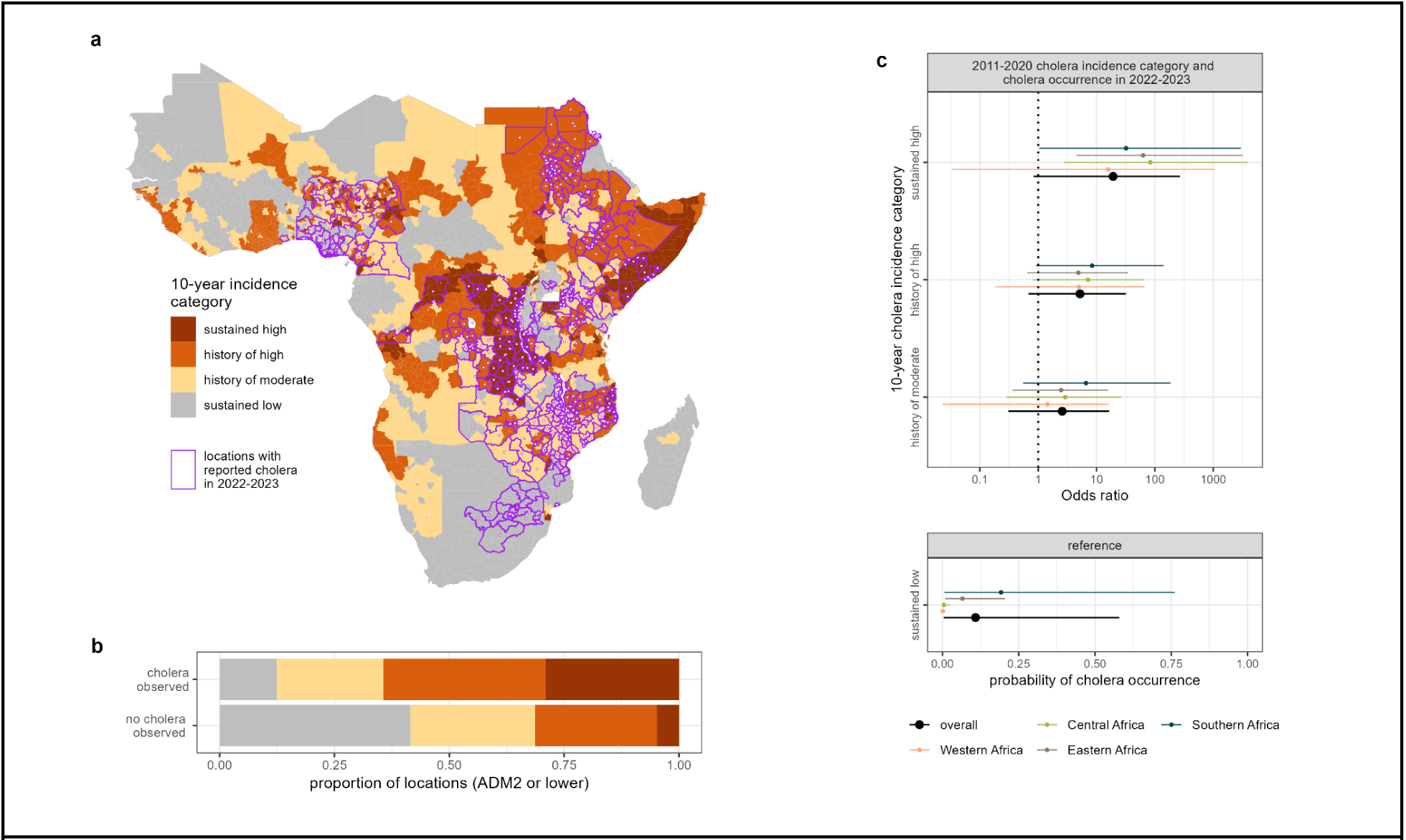
Associations between cholera occurrence in 2022-2023 and ten-year (2011-2020) cholera incidence categories in Africa. a) 2022-2023 cholera occurrence locations (purple borders with dots at location centroids) at the reported ADM scale overlaid on the 10-year incidence category map. b) Continent-wide distribution of 10-year incidence categories (colors) among ADM2 locations with reported cholera occurrence in 2022-2023. Locations with occurrence reported below ADM2 level were assigned to the corresponding ADM2 location. c) Pooled modeled estimates of the baseline probability of cholera occurrence in the sustained low incidence reference category (bottom panel) and odds ratios of reporting by 10-year incidence category relative to the reference (top panel, x-axis ticks are on the log scale but labels are on the natural scale), at the continent level (black) and by region (colors). Points indicate mean estimates and bars indicate 95% CrIs from 4000 HMC posterior draws.

Due to spatial clustering of areas with high burden, targeting interventions to ADM2 units based on incidence categories would enable reach to a greater proportion of cases than proportion of population targeted. For instance, assuming incidence categories are known (oracle), targeting the top 50 million highest burden population (roughly 5% of total population) would reach 29% (95% CrI: 28-31) of 2016-2020 cases, and 66% (95% CrI: 65-67) when targeting the top 100 million (10% of total population) (Fig 6), with similar or better yields in 2011-2015 (Figure S21). Using 2011-2015 patterns for planning 2016-2020 interventions (prospective) achieved yields similar to oracle targeting only for the top 50 million highest burden people and had significantly reduced intervention reach beyond. For example, targeting the top 100 million of the highest burden population in 2011-2015 would reach only 37% (95% CrI: 36-38) of 2016-2020 cholera cases. Further, we found that using more temporally-distal incidence categories for prioritization decreased reach of interventions applied to locations with cholera in 2022-2023. Targeting the top 100 million people based on 2016-2020 incidence categories reached 19% (95% CrI: 16-21) of the 2022-2023 cholera-affected population, versus 13% (95% CrI: 11-15) using 2011-2015 incidence categories, with an increasing difference in yield as more people were targeted (Fig 6). Using the 2011-2020 incidence categories for targeting performed similarly to targeting based on 2016-2020 incidence categories.

**Figure 6:**
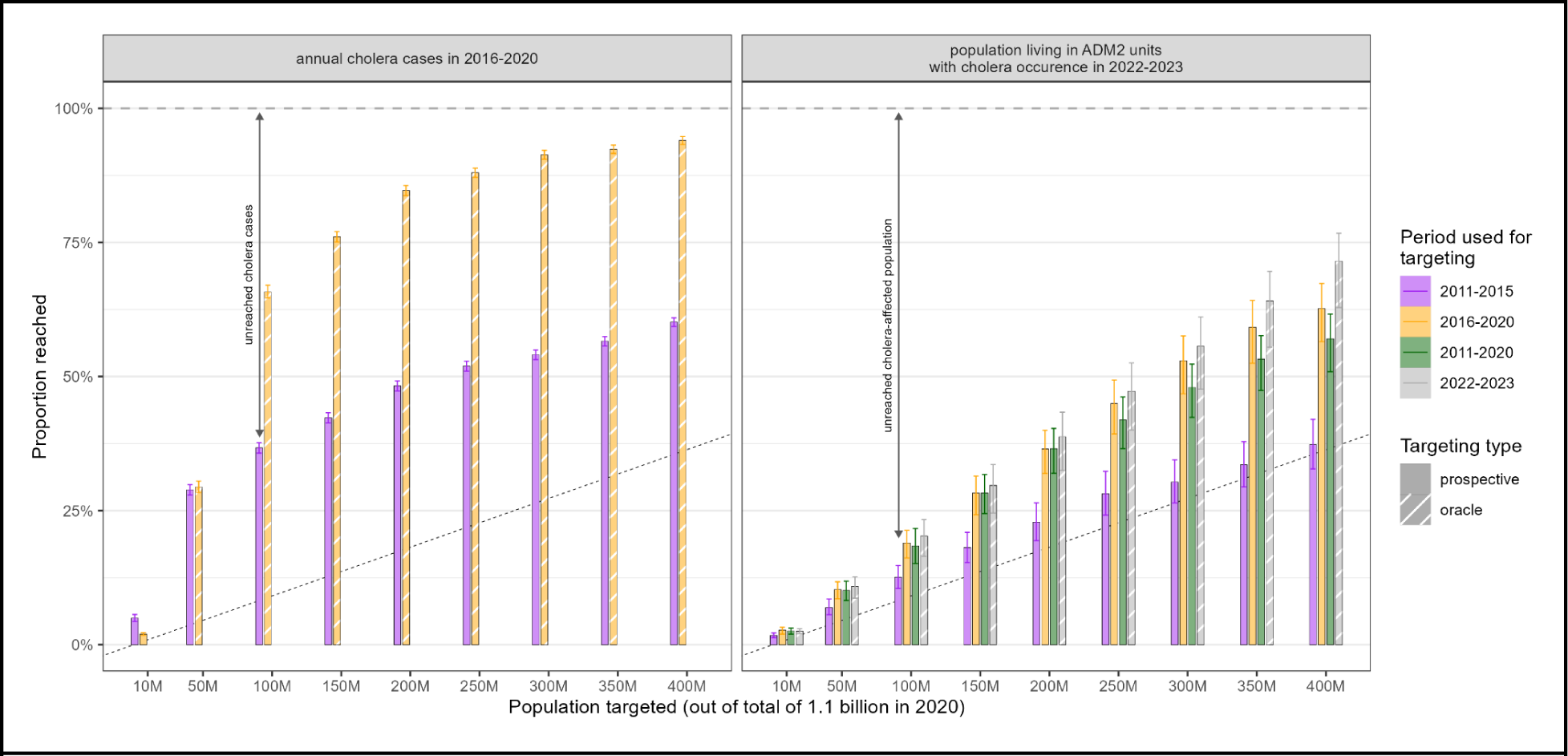
Potential reach of interventions prioritized by past cholera incidence categories. Proportion of 2016-2020 cases (left panel, y-axis) or population living in ADM2 units with cholera occurrence in 2022-2023 (right panel, y-axis) reached when prioritizing people living in ADM2 units (x-axis) by past incidence categories (“prospective” targeting, full bars) or burden in the concurrent period (“oracle” targeting, hashed bars). There is only one past incidence category (2011-2015) for the 2016-2020 period. There were three past incidence categories (2011-2015, 2016-2020, 2011-2020) for the 2022-2023 period. The horizontal dashed line marks 100% of cases or population reached. The diagonal dotted line indicates unit yield in population targeted (eg. targeting 10% of the population in Africa reaches 10% of cases in 2016-2020 or cholera-affected population in 2022-2023).

## Discussion

By developing high-resolution maps of cholera burden in Africa, we highlight challenges in making progress towards cholera control, given that cholera burden in the continent is both persistent and characterized by shifting patterns over time and space. We found that reported suspected cholera cases increased in 2016-2020 relative to 2011-2015 although the mean annual incidence rate remained stable across the two periods. Western Africa experienced a sharp decline in 2016-2020 relative to 2011-2015, which was offset by increases in cholera in Eastern Africa. We also saw geographic shifts at the subnational level in 16 cholera-affected countries. Nevertheless, consistent with previous analyses, cholera burden remained spatially concentrated, with only 18% of second-level administrative units having high cholera incidence in 2016-2020.

The spatial concentration of high cholera burden may be leveraged to set expectations about where interventions are critically needed and where they can be most efficiently deployed, particularly in the current context of limited resources and OCV supply ^7,15^. While everyone is entitled to safe water and sanitation, the enormous infrastructure investments that are needed will be most feasible and impactful if implemented in a staged, targeted approach that prioritizes rollout in high-risk populations. We estimated 82 million people living in areas with very high incidence and 296 million living in high incidence areas in 2016-2020. Taken with estimates between 2010 and 2015 and the spatial concentration of burden ^4^, these results identify a shortlist of potential spatial targets for high-impact infrastructure investments and demonstrate a large and sustained potential OCV demand that has consistently exceeded past and projected global production capacity (37-50 million doses in 2024) ^50^.

The public health impact of targeting interventions based on past incidence hinges on whether these spatial patterns remain stable over time. We find that some areas in Africa had sustained high incidence, while even more areas experienced it only sporadically. There were 105 million people living in second-level administrative units with high incidence throughout 2011 to 2020, who also tended towards higher odds of cholera in 2022 to 2023. Yet these 105 million constituted only a quarter of people that experienced any high incidence during the decade, and this figure is dwarfed by the number witnessing extreme shifts during this time (e.g., areas that were low incidence in 2011-2015 and high incidence in 2016-2020 were home to 135 million people). This instability in spatial patterns continued into 2022-2023, where we estimated an 11% probability of cholera among ADM2 units with sustained low incidence in 2011-2020 (driven mostly by areas in Southern Africa). Together, these results suggest areas in Africa with low incidence, even across a 10-year period, could still be vulnerable to large outbreaks upon *V. cholerae* introduction.

The spatiotemporal clustering we observed in cholera incidence is a double-edged sword for the effectiveness of geographic intervention targeting. While there are notable advantages to such a strategy, as targeting the top 10% of highest burden 2016-2020 populations would reach over 60% of cholera cases in that period, shifting spatial patterns mean that this yield decreases significantly when using past patterns to target future interventions (e.g., targeting the top 10% of 2011-2015 highest burden populations would reach only 37% of 2016-2020 cases). This suggests that when countries identify priority areas for multisectoral interventions (PAMIs) for national cholera planning following GTFCC recommendations ^51^, the robustness of PAMI selections should be evaluated across multiple time ranges and consider changing epidemiologic context and risk factors. Nevertheless, our results suggest that targeting populations with recent very high incidence (≥100 per 100,000 population) or high incidence over sustained periods (≥10 per 100,000 population over 10 years) has the best potential to maximize the efficiency of cholera intervention reach.

Interpretation of the magnitude and spatial distribution of these maps are limited by the underlying data, which comprises medically-attended suspected cholera across multiple case definitions and varying adaptively by transmission setting and location ^52^. In reality, we know that *V. cholerae* may account on average for only half of suspected cases, with high variability across settings, and we expect to see improvements in the estimation of true cholera incidence in the coming years with expanding RDT usage and clearer testing guidance ^9,52^. Nevertheless, we believe our maps and modeling methodology improve upon previous estimates of cholera burden in Africa due to substantial enhancements to spatial data coverage, data processing, and modeling fidelity ^4^.

The stability in continent-wide cholera incidence rates from 2011-2020 may be disheartening to those who know how much effort has been invested in cholera control during this period. It is important to note, however, that improvements in surveillance may confound the interpretation of cholera burden changes in our analysis. Notably, Ethiopia and Sudan had limited data available in 2011-2015 and high reported burden in 2016-2020, suggesting that apparent burden increases could be due largely to changes in reporting and data availability. These challenges highlight that continued efforts to sustain high-quality surveillance are needed to better characterize long-term changes in cholera epidemiology.

The instability in cholera burden we observed in our analysis may be exacerbated by climatic and sociopolitical factors, and multisectoral control efforts and research must jointly accelerate to offset a possible increase in cholera vulnerability. Our continent-wide analysis pushes us in this direction, as it provides important context and supplements country analyses to identify priority intervention areas, which incorporate local knowledge of risk factors and surveillance gaps at more resolved spatial and temporal scales ^53–64^. Regular high-level mapping analyses such as this are therefore critical to keeping the global cholera response relevant and to tracking progress towards disease control goals.

## Data Availability

https://cholera-taxonomy.middle-distance.com/

https://github.com/HopkinsIDD/cholera-mapping-pipeline/releases/tag/v1.0

## Additional files

Supplementary figures (Figures S1-S21), tables (Tables S1-S7), and methods are available in the Supporting Material (SM).

## Author contributions

ASA, ECL, and JL conceptualized the study and acquired the funding. ASA, CA, ECL, JK, MD, RD, and QZ curated the data. ECL, JK, JPS, KZ, and QZ did the formal analysis. AD, ASA, ECL, JK, JPS, KZ, and QZ performed the investigation. AD, ASA, ECL, JK, JL, and JPS designed the methodology. ASA and ECL administered the project and provisioned the resources. ECL, JPS, JK, KZ, and QZ developed the software. ECL supervised the project. AD, ASA, ECL, JPS, QZ, JPML, RC, PWO, GB, AN, LE, NFM, EWO, SY, and FK validated the results. JPS and QZ developed the visualizations. JPS and ECL wrote the original draft. All authors reviewed and edited the draft.

## Acknowledgments

The authors would like to thank the numerous institutions and organizations that contributed surveillance data, including the World Health Organization, UNICEF, Epicentre, the DOVE Project, and many ministries of health. They would also like to acknowledge the staff programmer Pengcheng Fang, and former team members that have contributed to data entry into the Cholera Taxonomy database over the years. Early feedback on this work was provided by the WHO Cholera Team, the GTFCC Country Support Platform, GTFCC OCV Working Group, GTFCC Surveillance Working Group, and cholera focal points at ministries of health and WHO country and regional offices. Modeling was carried out at the Advanced Research Computing at Hopkins (ARCH) core facility (rockfish.jhu.edu), which is supported by the National Science Foundation (NSF) grant number OAC 1920103.

