## Supporting Material for "A decade of shifting cholera burden in Africa and its implications for control: a statistical mapping analysis"


**Supplementary Figures** 2
**Supplementary Tables** 17
**Supplementary Material** 26
**Cholera data sources and data processing** 26
Cholera data collection and data template 26
Temporal aggregation 26
Identifying time-censored observations 27
National-level data imputation 27
Geographic linkage of observations to modeling grid 28
**Incidence modeling framework** 28
Base model 28
Prior on the spatial random effect (s) 29
Prior on the temporal random effect (t) 30
Expansion for partial-year observations 30
Expansion for overdispersed observation data 31
Complete standard model formulation 31
Model formulation without spatial autoregressive term 32
Model selection and model formulation with non-mixture prior 32
Incidence modeling post-processing 32
**Analysis of 2022-2023 cholera outbreak occurrence** 34
Extraction and spatial linkage of cholera occurrence data 34
Base statistical model 34
Adding higher administrative unit level observations 35
Hierarchical country and region-level priors 35
Model priors and hyperpriors 36
Assessing intervention reach when prioritizing targets by cholera incidence 36
**References** 36

#### Supplementary Figures

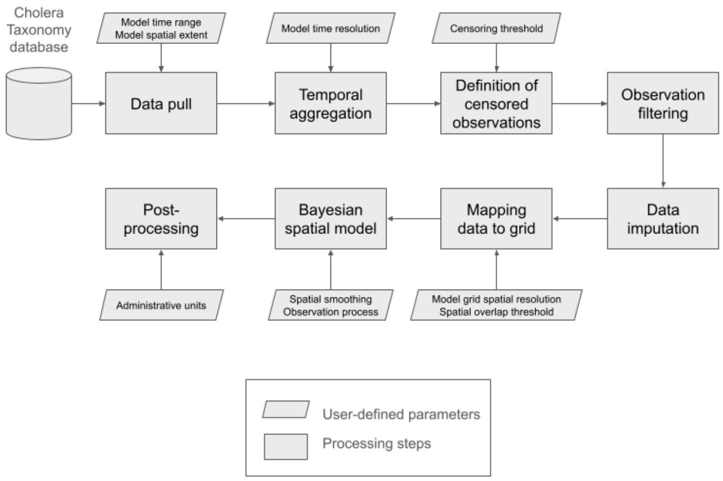


**Figure S1.** Flowchart summarizing the data processing and modeling pipeline and the corresponding user-defined inputs.

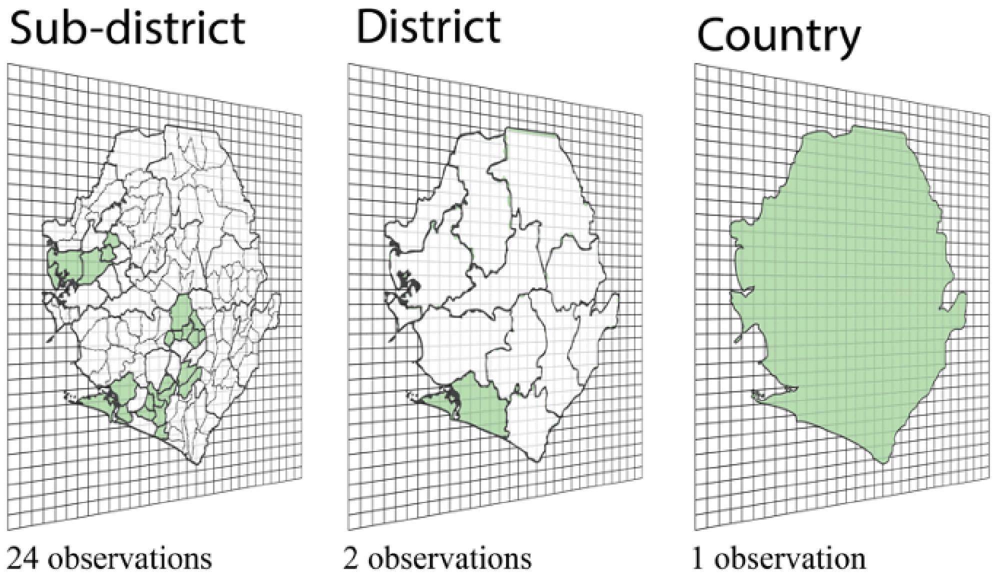


**Figure S2.** Conceptual diagram of how observations are mapped to the spatial modeling grid. Observations linked to geographic shapefiles (green) at different spatial scales are overlaid onto a 20 km by 20 km grid. Grid cells whose centroids overlap with the shapefile are linked to the observation.

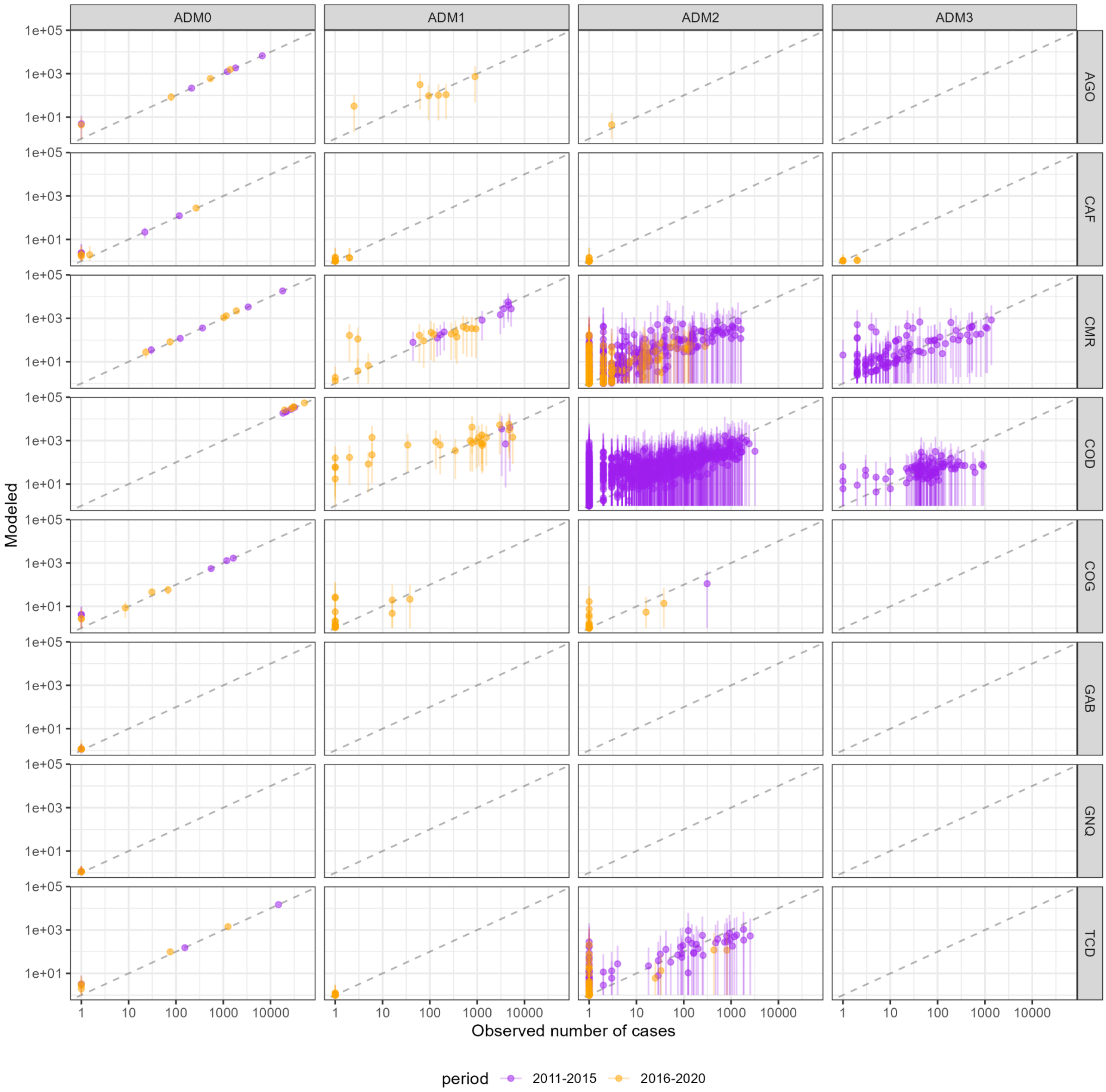


**Figure S3.** Scatterplot of full-year observations versus fitted values by administrative unit levels 0 to 3 for countries in Central Africa (log-scale). Dots and lines represent the mean and 95% CrI and colors indicate observations in different periods. Points falling along the 1:1 diagonal black dashed line indicates alignment between modeled and observed cases.

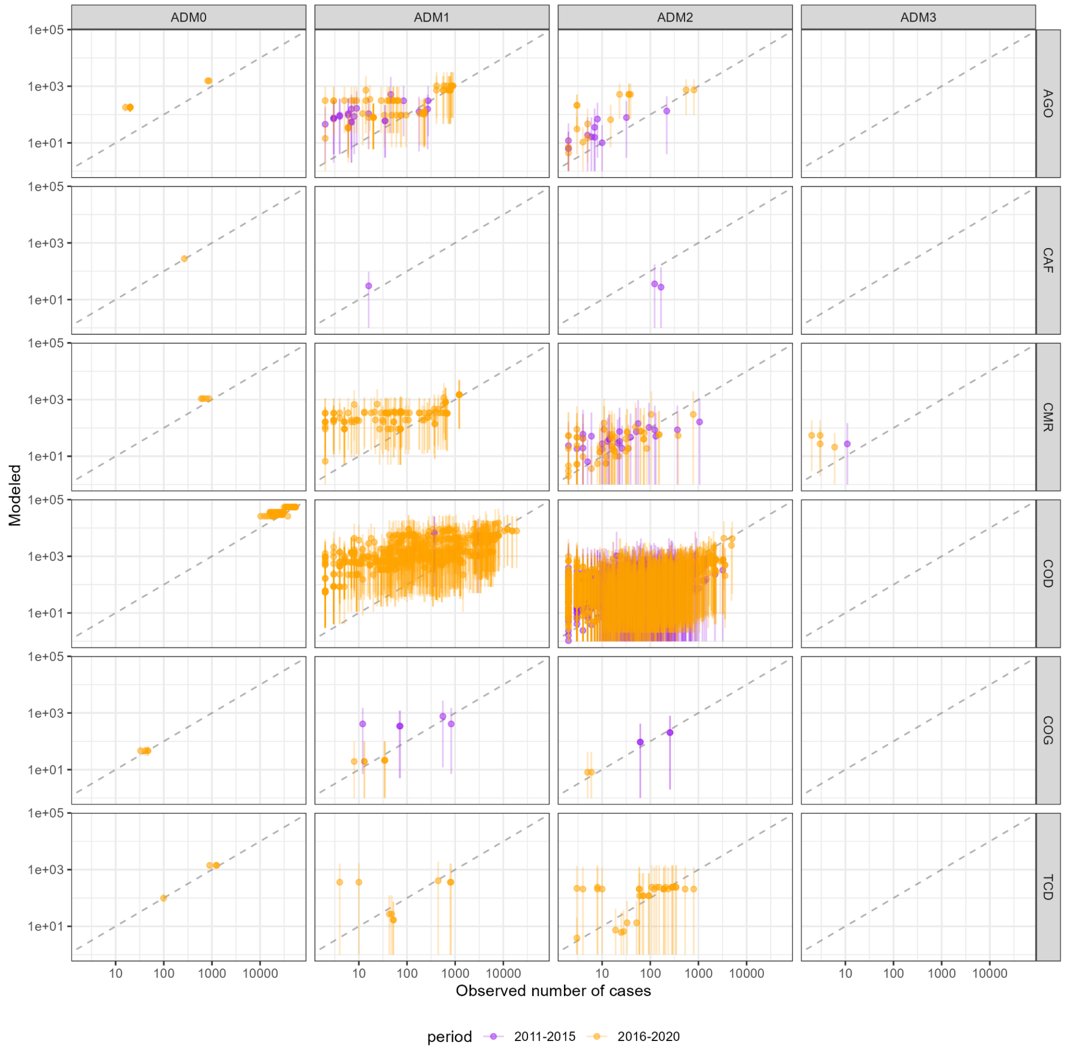


**Figure S4.** Scatterplot of partial-year observations versus fitted values by administrative unit levels 0 to 3 for countries in Central Africa. Dots and lines represent the mean and 95% CrI and colors indicate observations in different periods. Partial-year observations were treated as right-censored in the model likelihood. Models that fit well should have fitted values (y-axis) at or above their corresponding model observation values (x-axis).

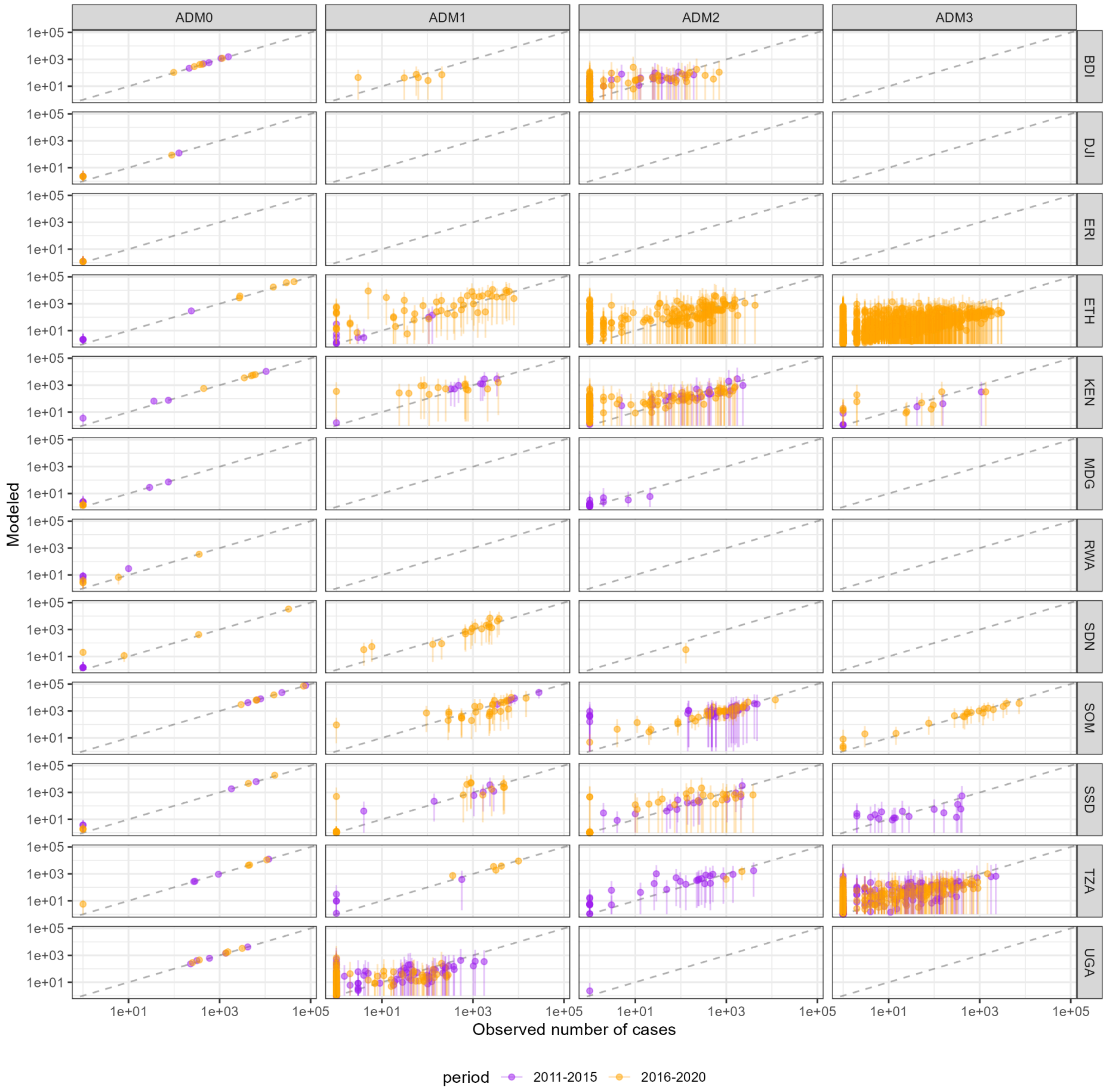


**Figure S5.** Scatterplot of full-year observations versus fitted values by administrative unit levels 0 to 3 for countries in Eastern Africa. Legend as in Figure S3.

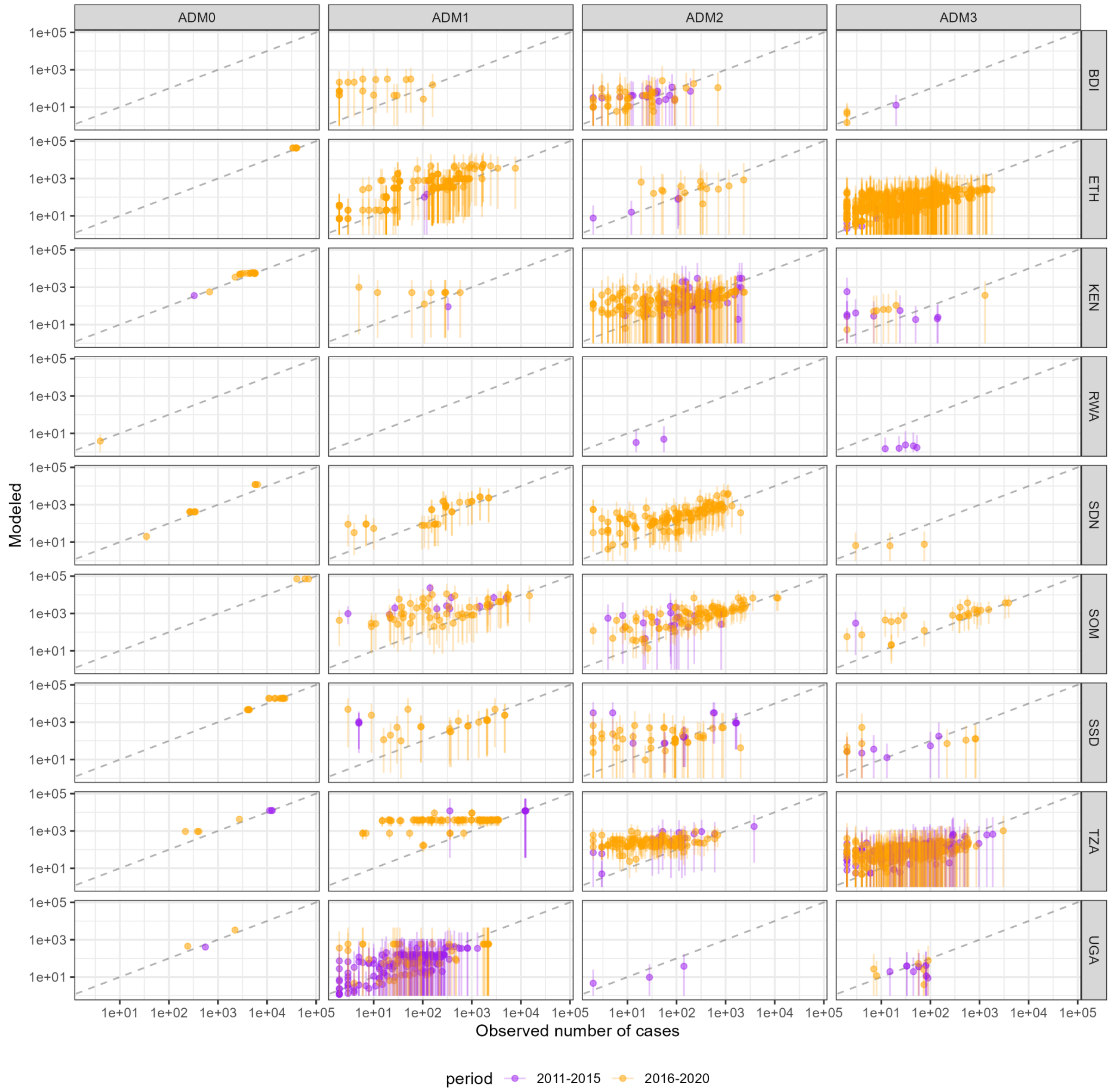


**Figure S6.** Scatterplot of partial-year observations versus fitted values by administrative unit levels 0 to 3 for countries in Eastern Africa. Legend as in Figure S4.

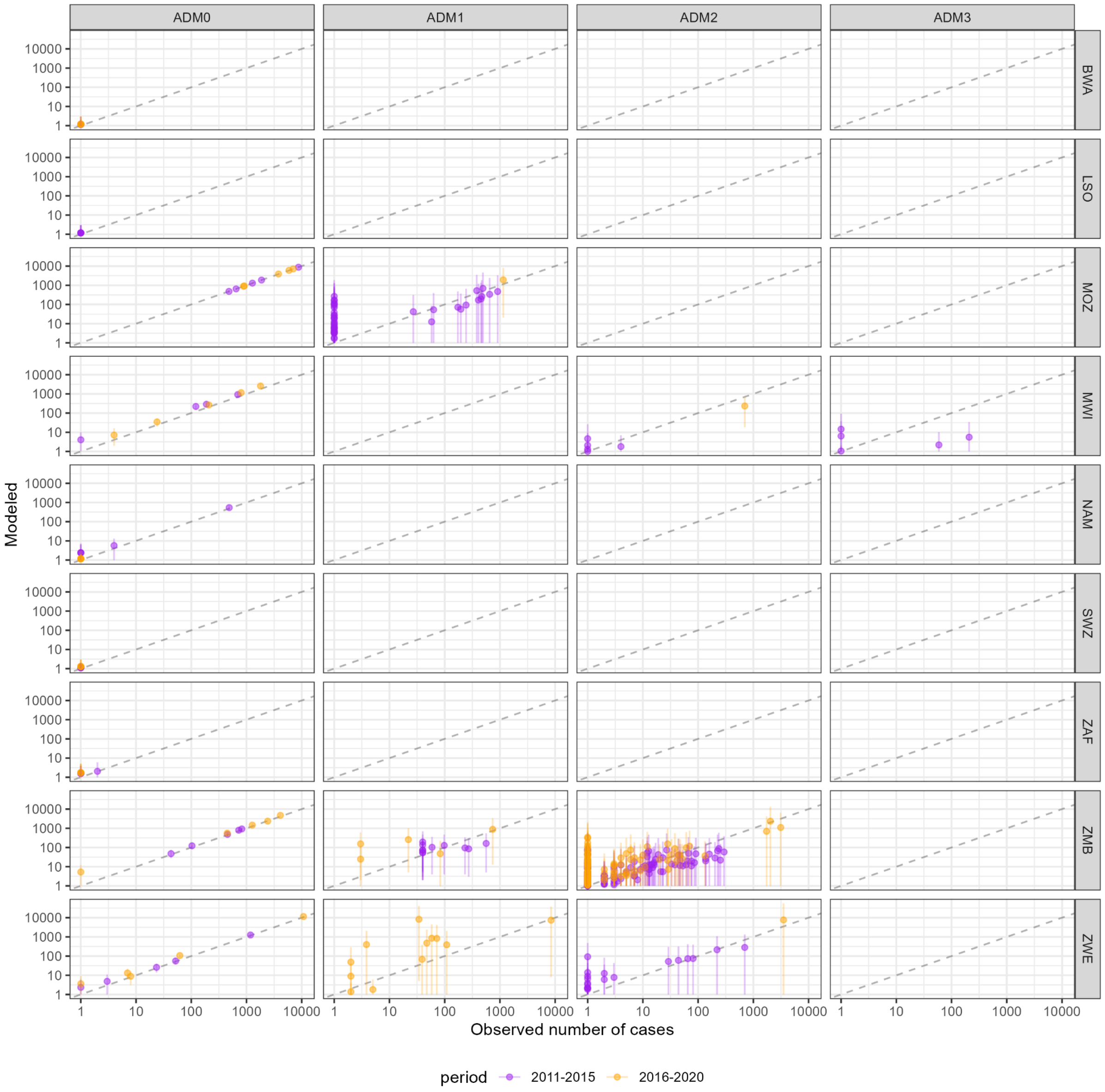


**Figure S7.** Scatterplot of full-year observations versus fitted values by administrative unit levels 0 to 3 for countries in Southern Africa. Legend as in Figure S3.

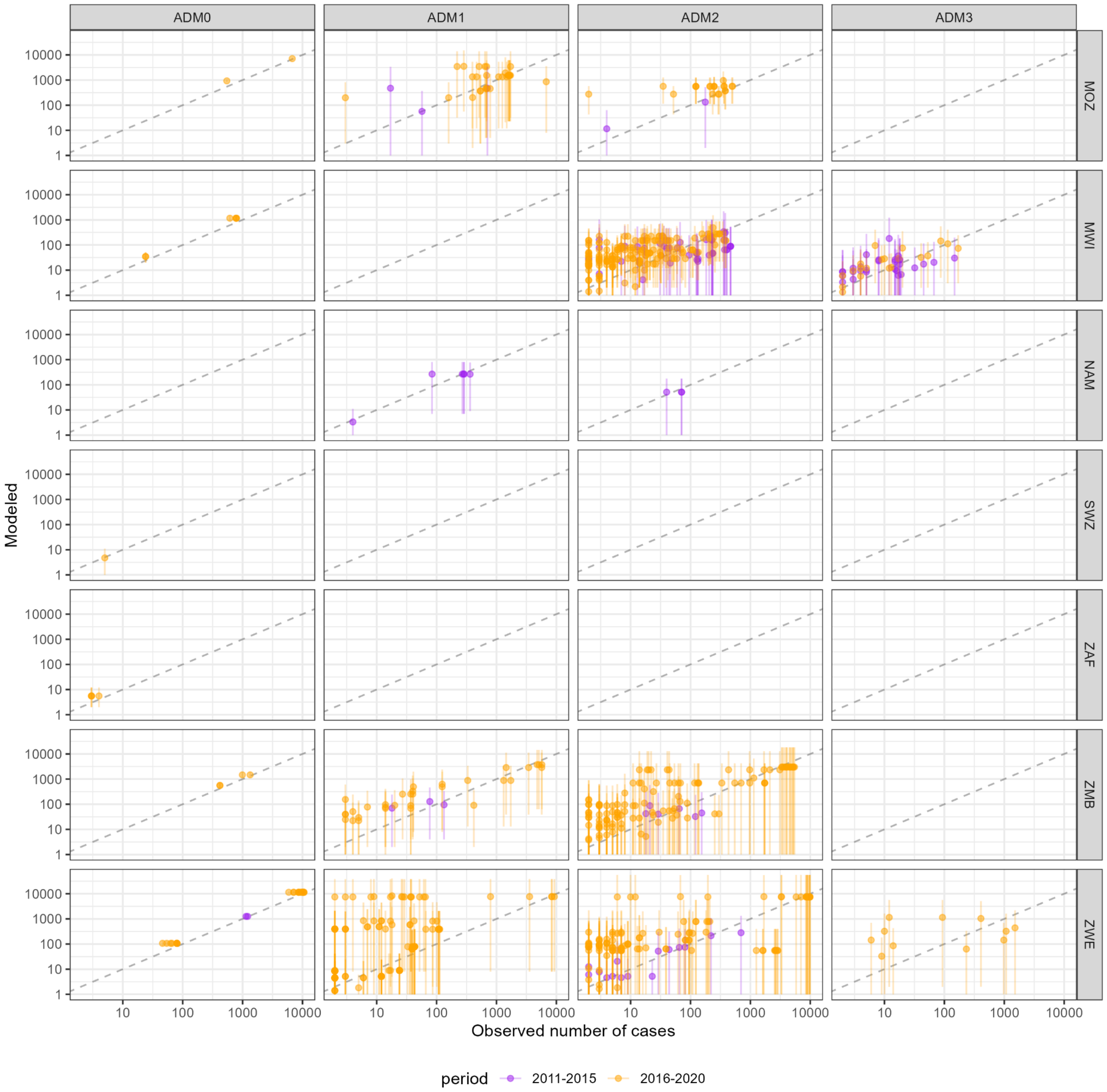


**Figure S8.** Scatterplot of partial-year observations versus fitted values by administrative unit levels 0 to 3 for countries in Southern Africa. Legend as in Figure S4.

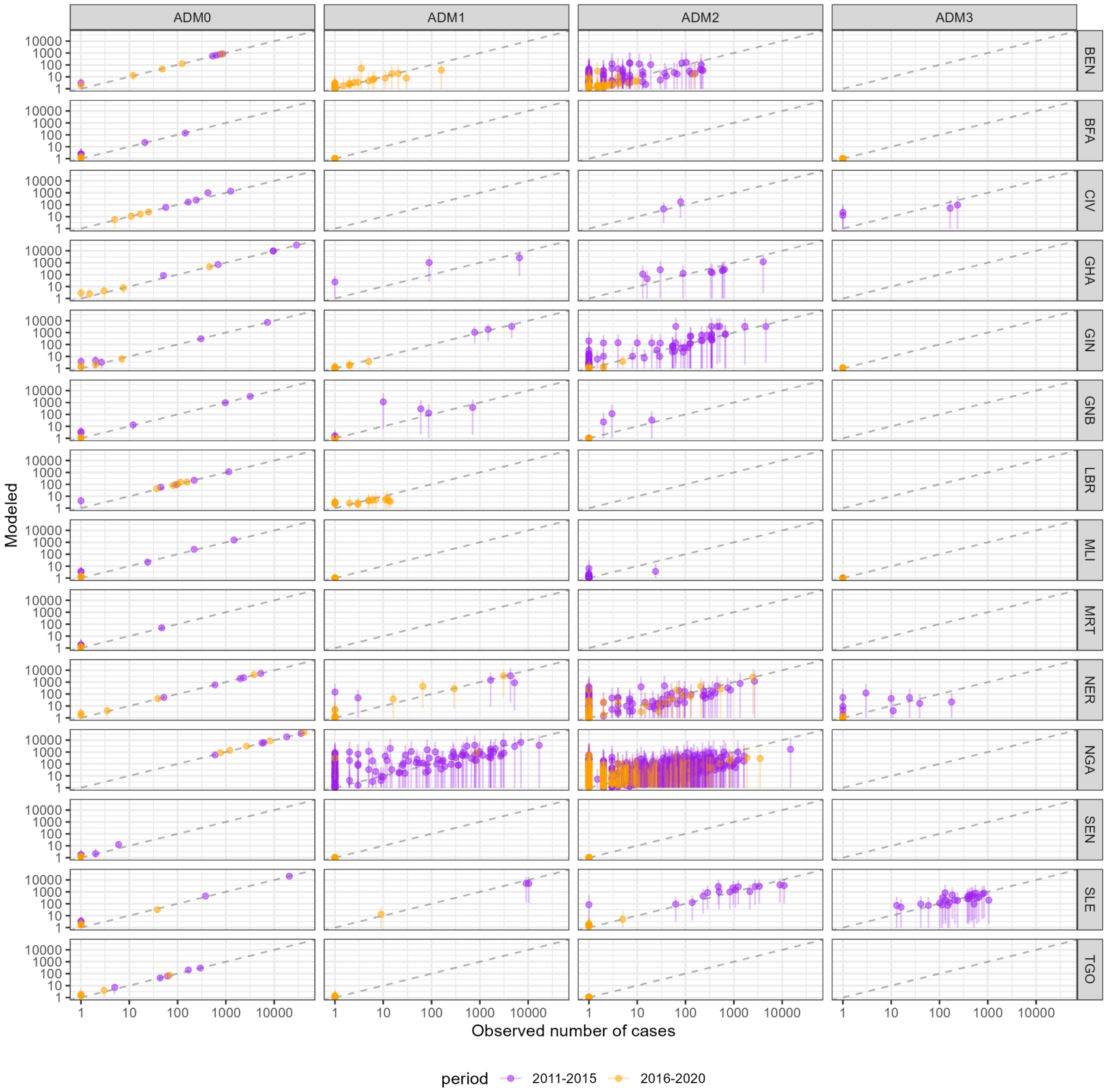


**Figure S9.** Scatterplot of full-year observations versus fitted values by administrative unit levels 0 to 3 for countries in Western Africa. Legend as in Figure S3.

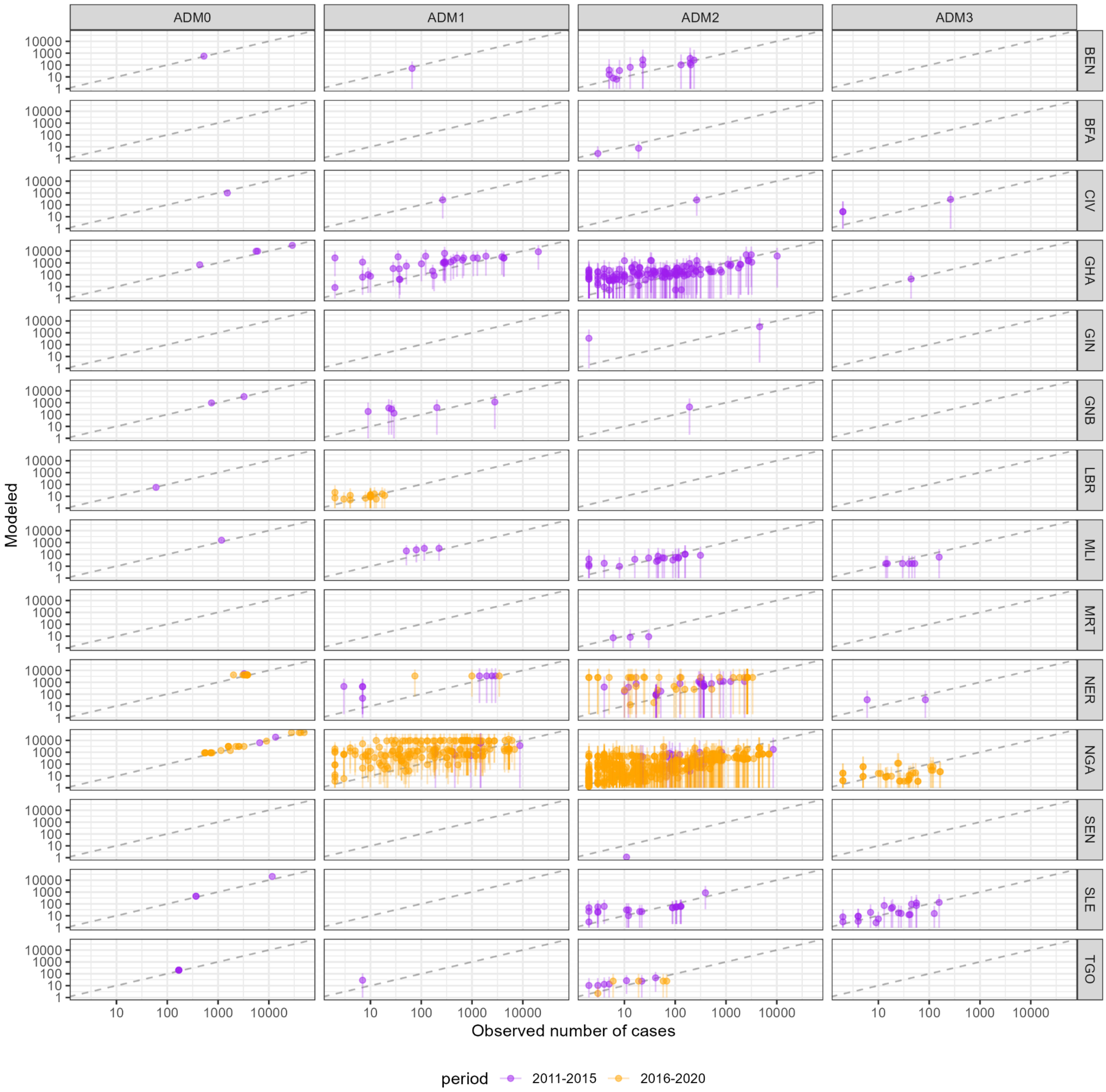


**Figure S10.** Scatterplot of partial-year observations versus fitted values by administrative unit levels 0 to 3 for countries in Western Africa. Legend as in Figure 4.

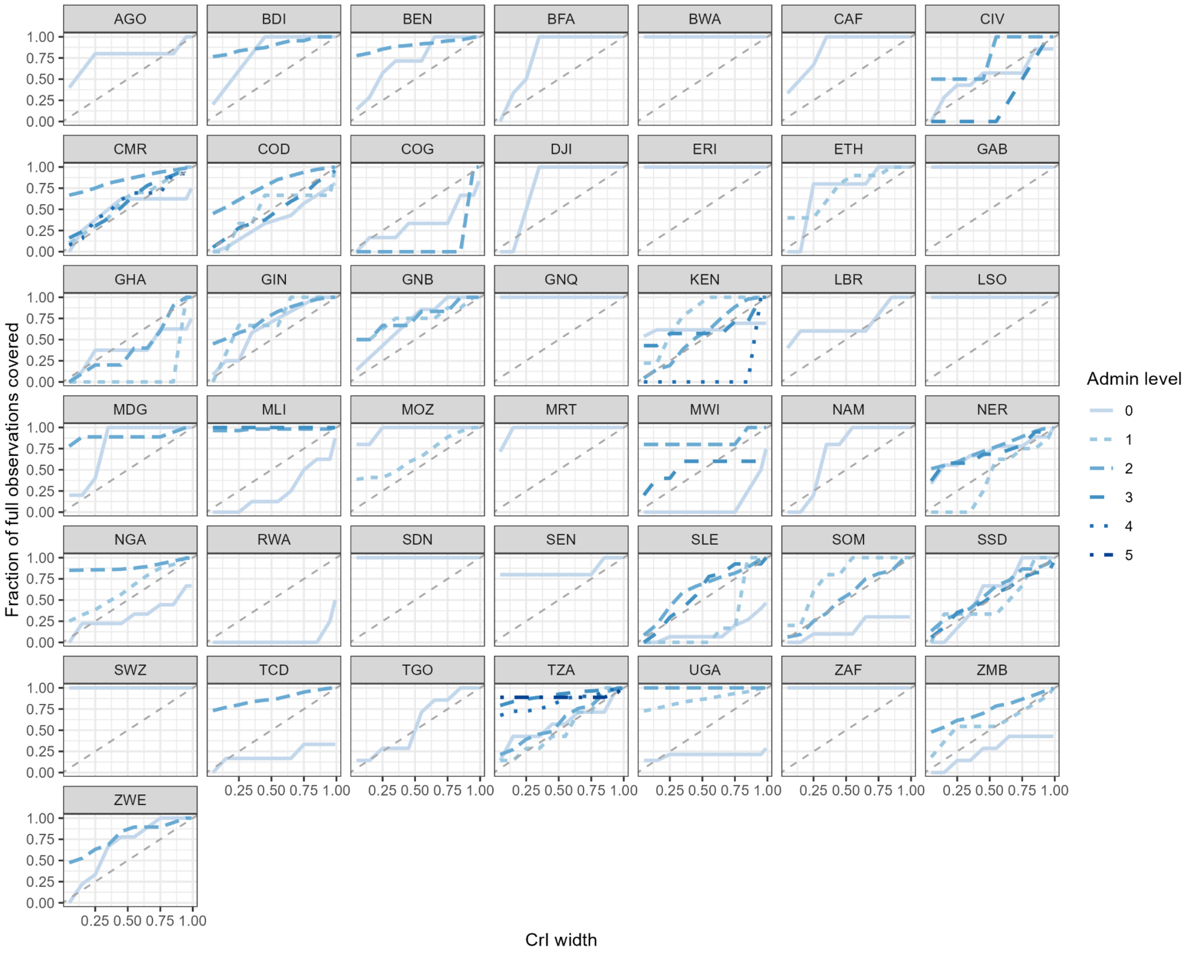


**Figure S11.** Coverage of full-year model observations by administrative unit level versus the credible interval width for 2011-2015 models. This figure indicates the appropriateness of the width of the fitted credible intervals based on the spread of the observation data. In a model that is neither overfit (with an interval that is too narrow) nor underfit (with an interval that is too wide), the credible interval width should roughly match the fraction of observations covered by the interval. For example, at the 50% CrI (x-axis), 50% of full observations (y-axis) should fall within the interval. Note that administrative unit level 0 (country-level) coverage is not comparable to that of other levels because the model fit is more restricted for these observations.

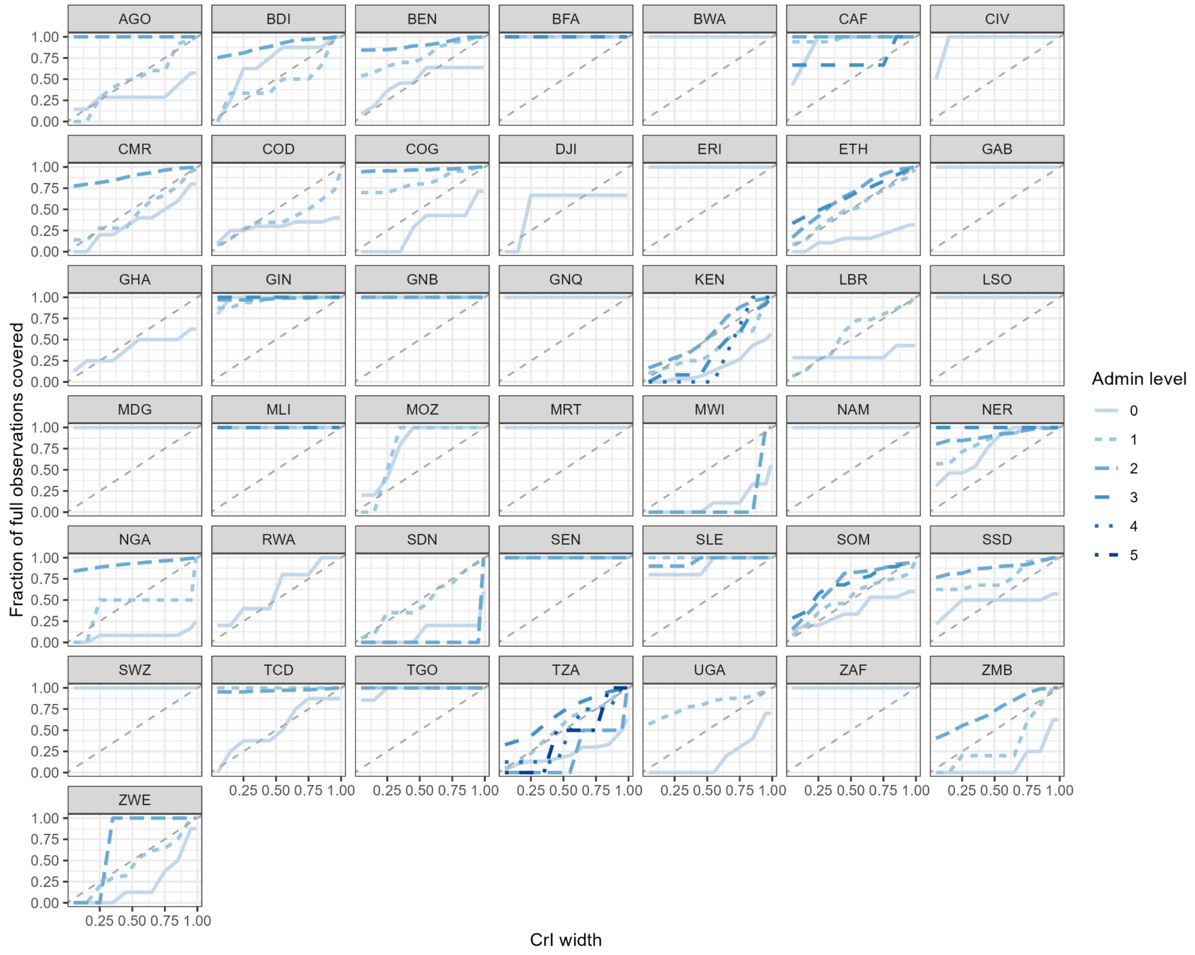


**Figure S12.** Coverage of full-year model observations by administrative unit level versus the credible interval width for 2016-2020 models. Legend as in Figure S11.

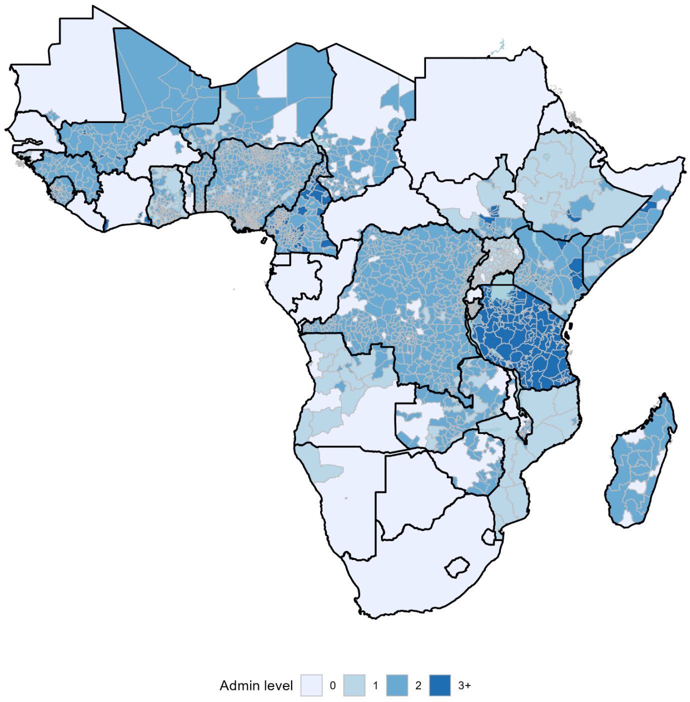


**Figure S13.** Spatial coverage of cholera observation data for the 2011-2015 period. Colors represent the smallest administrative unit level with at least one observation available during this period. For example, any spatial area with a yellow fill was covered only by an administrative unit level 0 (country-level) observation, while any spatial area with a red fill was covered by at least one administrative unit level 3 (subdistrict-level) observation. Only modeled countries are shown.

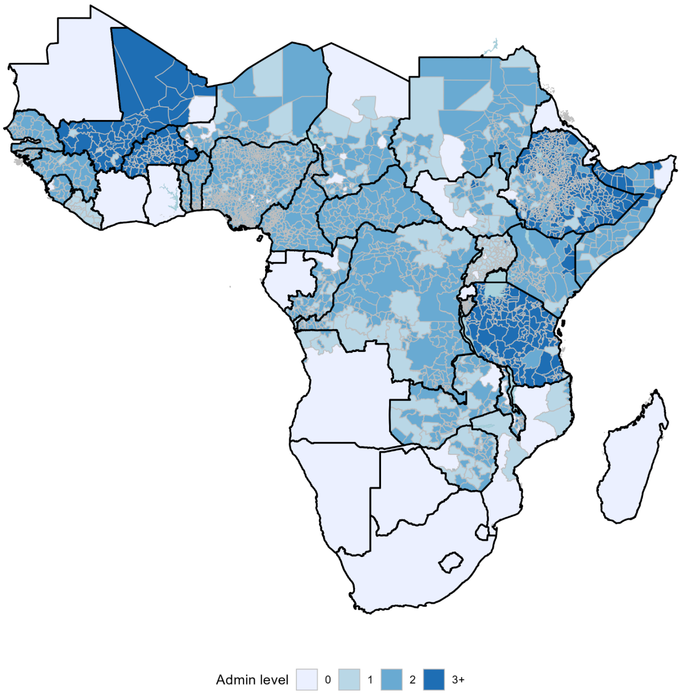


**Figure S14.** Spatial coverage of cholera observation data for the 2016-2020 period. Legend as in Figure S13.

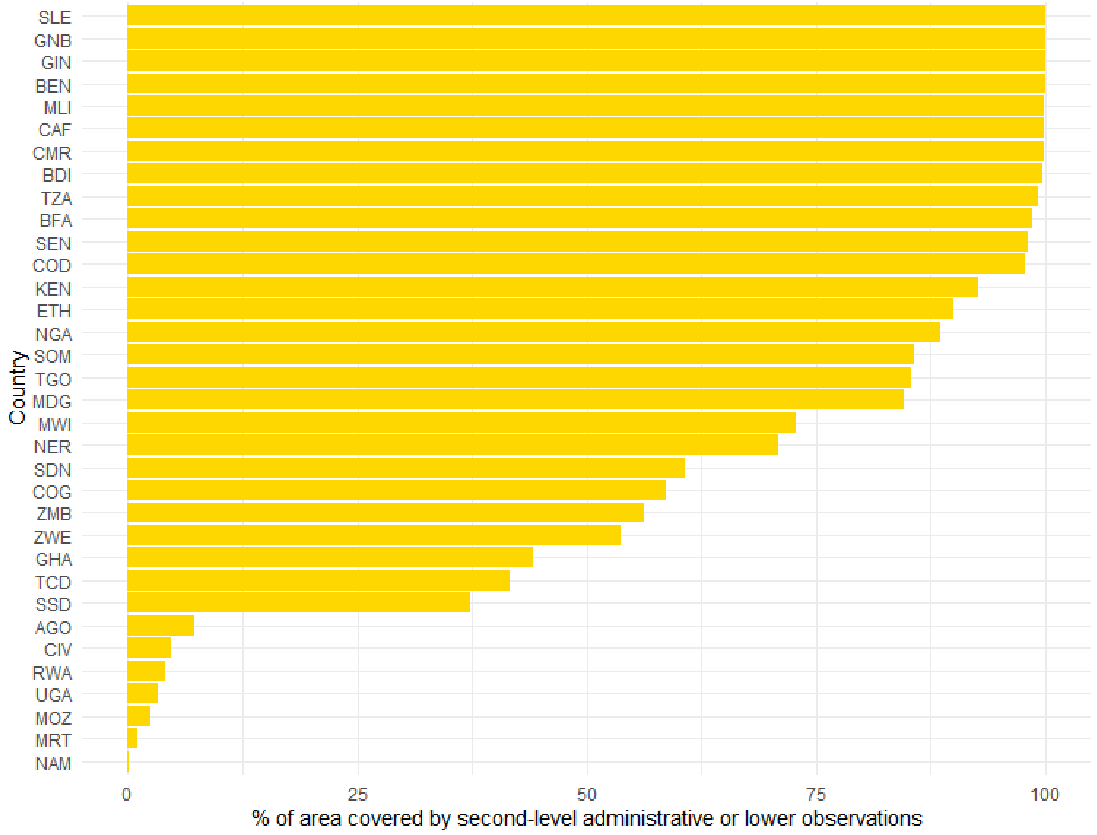


**Figure S15.** Percent of spatial area in the country that is covered by administrative unit level 2 or lower observations in at least one year from 2011-2020. Countries are displayed in descending rank order.

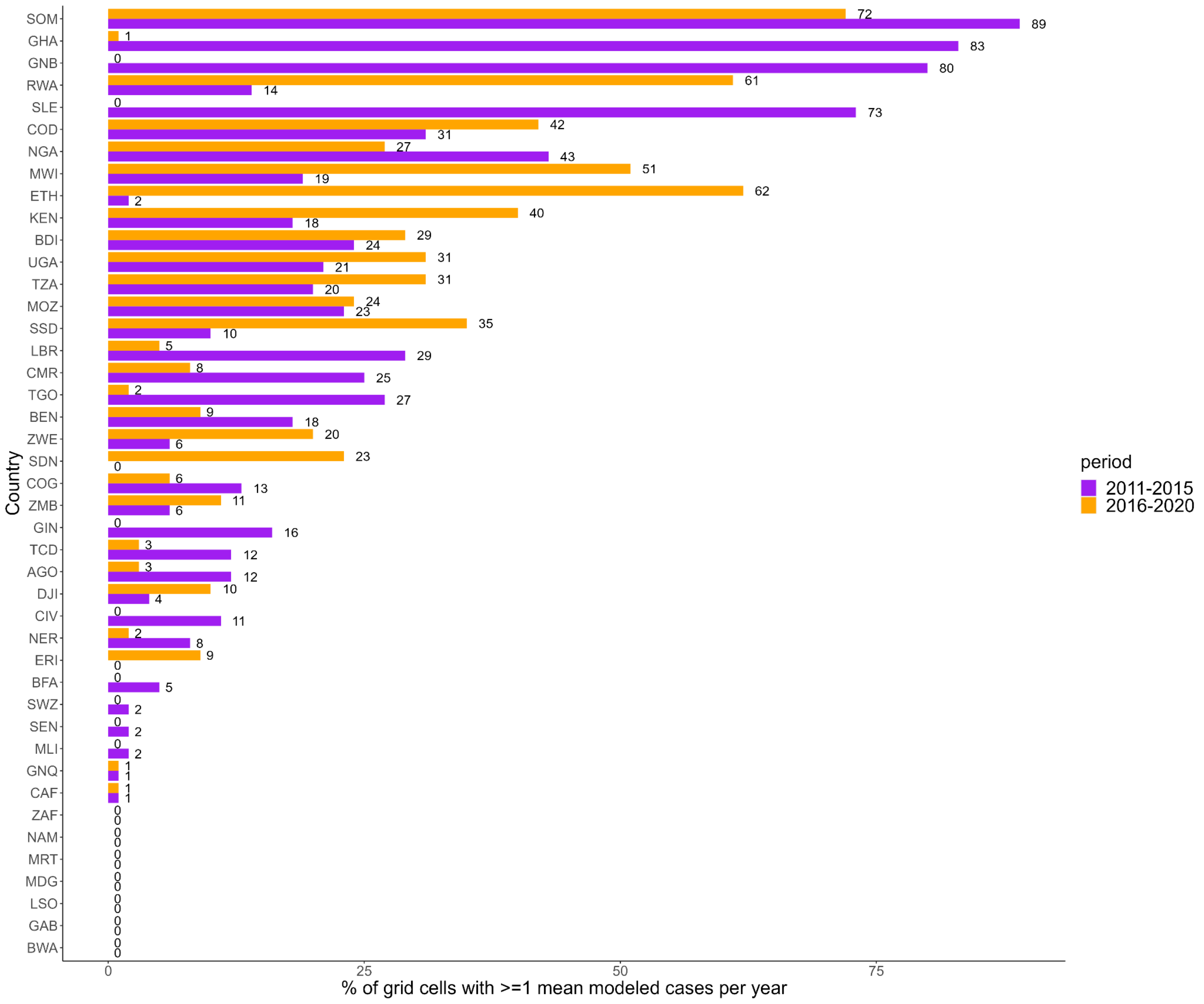


**Figure S16.** Percent of 20 km by 20 km grid cells in the country spatial modeling grid with at least 1 suspected case estimated per year, by country and period. Colors differentiate the two periods and labels indicate the percent value.

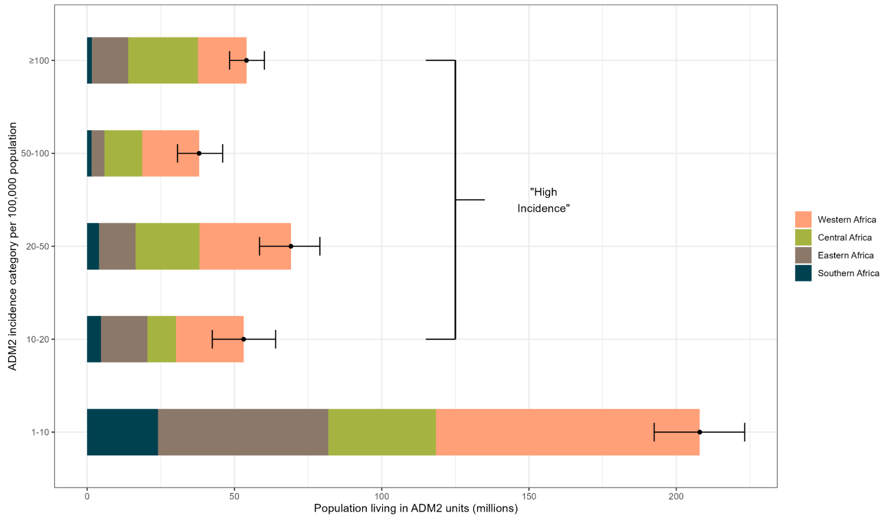


**Figure S17.** Population living in areas by incidence category and region in 2011-2015. Mean and 95% CrI for ADM2 populations living in a given incidence category (per 100,000 population) across Africa. Regional population contributions are indicated by fill colors.

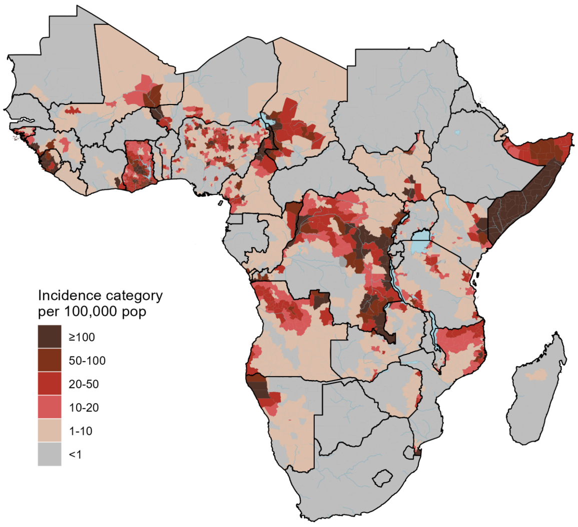


**Figure S18.** Continent-wide map showing assignment of incidence categories to ADM2 units for 2011-2015. ADM2 units were assigned to an incidence category if 50% of posterior draws classified the ADM2 unit to the assigned color of incidence category or above. ADM2 units in gray had an incidence category of <1 per 100,000 population. Only modeled countries are displayed in the map.

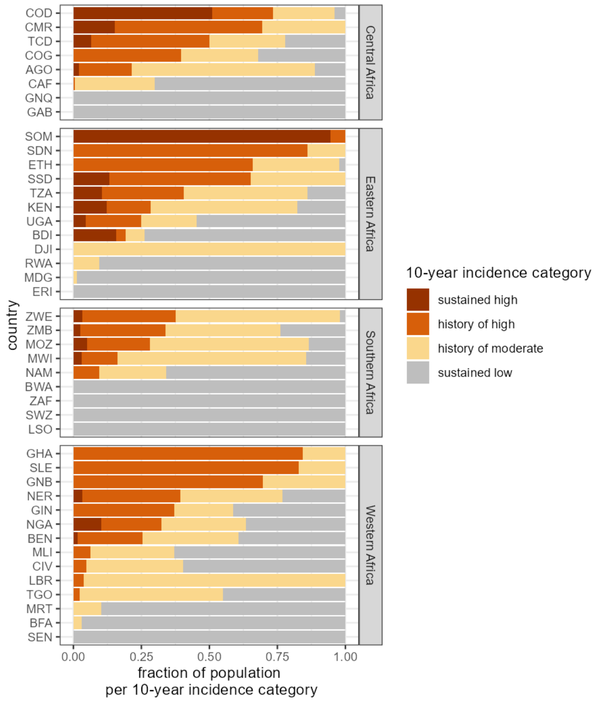


**Figure S19.** Distribution of population living in ADM2 units in each 10-year incidence category by country. Countries are grouped by region and displayed in descending order by the sum of the population fraction in the sustained and history of high-incidence categories.

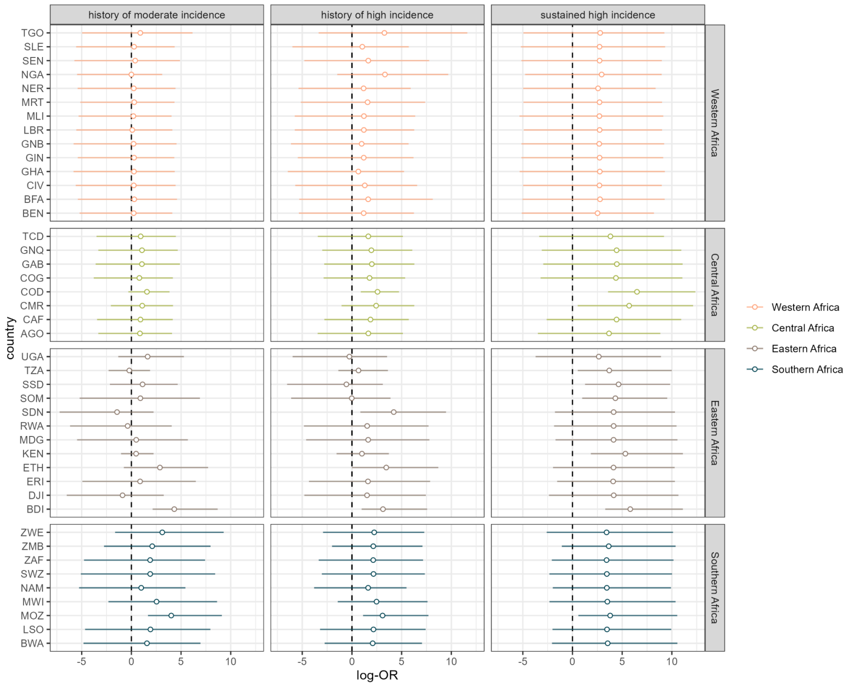


**Figure S20.** Log-odds ratios of reporting cholera occurrence in the post-2020 period by 10-year incidence category relative to the baseline probability of cholera occurrence in the sustained low incidence reference category by country. Countries are grouped by region in facets and by color. Bars indicate the 95% CrIs from 4000 HMC posterior draws.

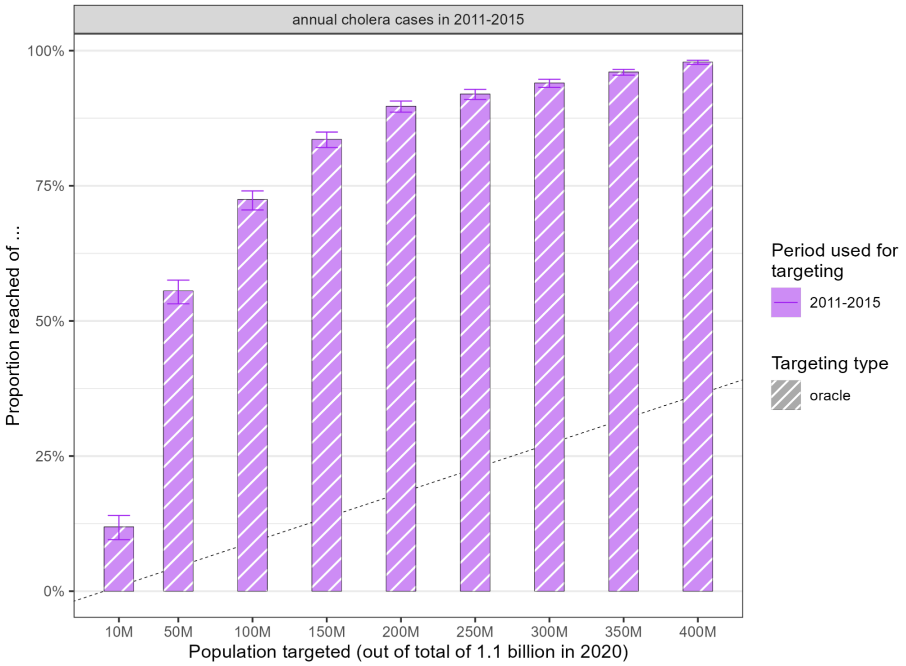


**Figure S21.** Proportion of 2011-2015 cases (y-axis) reached when prioritizing people living in ADM2 units (x-axis) by 2011-2015 incidence categories (“oracle” targeting).

### Supplementary Tables

**Table S1.** Comprehensive list of suspected case definitions and number of unique data sources using this definition.

| No. | Case definition | Number of sources |
|---|---|---|
| 1 | Any person presents with or dies from acute watery diarrhoea. | 491 |
| 2 | Unknown (*The document indicates that the case reports represent cholera, but no specific case definition is provided.*) | 230 |
| 3 | In a non-epidemic setting, a patient aged 5 years or more develops severe dehydration or dies from acute watery diarrhoea. | 26 |
| 4 | A patient aged 2 years or more develops acute watery diarrhoea, with or without vomiting. | 21 |
| 5 | In an epidemic setting, a patient aged 2 years or more develops acute watery diarrhoea, with or without vomiting. | 20 |
| 6 | Any person presents with acute watery diarrhoea. | 3 |
| 7 | In a non-epidemic setting, any patient presents with severe dehydration or dies from acute watery diarrhoea. | 2 |
| 8 | A patient aged 5 years or more develops acute watery diarrhoea and severe dehydration, with or without vomiting. In an epidemic setting, a patient aged 2 years or more develops acute watery diarrhoea, with or without vomiting. | 1 |
| 9 | A patient aged 5 years or more develops acute watery diarrhoea, with or without vomiting and was hospitalised for at least 1 night and treated with intravenous fluids. | 1 |
| 10 | A patient aged 5 years or more develops acute watery diarrhoea, with or without vomiting. | 1 |
| 11 | A patient aged 5 years or more develops severe dehydration or dies from 3 or more acute watery stools per day, with or without vomiting. A patient aged 2-4 years develops severe dehydration or dies from acute watery diarrhoea, with or without vomiting. | 1 |
| 12 | Any patient develops acute watery non-bloody diarrhoea with more than 3 liquid stools in a day. | 1 |
| 13 | Any person aged 2 years or more presents with acute watery diarrhoea and severe dehydration or dies from acute watery diarrhoea. | 1 |
| 14 | Any person presents with or dies from acute non-bloody watery diarrhoea with more than three liquid stools per day. | 1 |
| 15 | At the community level, any person presents with or dies from acute watery diarrhoea. At the health facility level and in a non-epidemic setting, a patient aged 5 years or more develops acute watery diarrhoea, with or without vomiting. At the health facility level and in an epidemic setting, a patient aged 2 years or more develops acute watery diarrhoea, with or without vomiting. | 1 |
| 16 | In an endemic setting, any person presents with acute watery diarrhoea. | 1 |
| 17 | In a non-epidemic setting, a patient aged 2 years or more develops acute watery diarrhoea and severe dehydration or death, with or without vomiting. In an epidemic setting, any person presents with or dies from acute watery diarrhoea. | 1 |
| 18 | In a non-epidemic setting, a patient aged 5 years or more develops severe dehydration or dies from acute watery diarrhoea. In an epidemic setting, a patient aged 5 years or | 1 |

| | | |
|---|---|---|
| | more develops acute watery diarrhoea. | |
| 19 | In a non-endemic setting, a patient aged 5 years or more develops acute watery diarrhoea with severe dehydration or death, with or without vomiting. In an epidemic setting, any person presents 3 or more liquid stools with or without vomiting for the pas 24 hours. | 1 |
| 20 | In an non-epidemic setting, a patient aged 5 years or more develops acute watery diarrhoea and severe dehydration, with or without vomiting. In an epidemic setting, a patient aged 2 years or more develops acute watery diarrhoea, with or without vomiting. | 1 |

**Table S2.** Summary of model observations by period. This table represents observation counts after data processing steps including temporal aggregation, but it excludes imputed national-level observations.

| Period | Data sources | Countries | Locations | Administ rative levels | Observations | National Observati ons | Subnation al observatio ns |
|---|---|---|---|---|---|---|---|
| 2011-15 | 291 | 43 | 2,944 | 6 | 14,872 | 276 | 14,596 |
| 2016-20 | 622 | 43 | 3,473 | 7 | 15,230 | 587 | 14,643 |
| 2011-20 | 807 | 43 | 4,574 | 7 | 30,102 | 863 | 29,239 |

**Table S3.** List of statistical model structures and data processing settings for each country and time period. Blank cells indicate that there was either no data processing deviation or no non-standard settings.

| Country (ISO3 code) | Period | Model | Data processing deviation, if any | Reasons for non-standard settings |
|---|---|---|---|---|
| Angola (AGO) | 2011-2015 | Non-mixture prior | - | Limited subnational data |
| | 2016-2020 | Non-mixture prior | - | Limited subnational data |
| Burundi (BDI) | 2011-2015 | Standard | - | - |
| | 2016-2020 | Standard | - | - |
| Benin (BEN) | 2011-2015 | Standard | - | - |
| | 2016-2020 | Standard | - | - |
| Burkina Faso (BFA) | 2011-2015 | Non-mixture prior | - | Limited subnational data |
| | 2016-2020 | No spatial autoregressive term | - | Zero cases |
| Botswana (BWA) | 2011-2015 | No spatial autoregressive term | - | Zero cases |
| | 2016-2020 | No spatial autoregressive term | - | Zero cases |
| Central African Republic (CAF) | 2011-2015 | Non-mixture prior | - | Limited subnational data |
| | 2016-2020 | Non-mixture prior | - | Limited subnational data |
| Côte d'Ivoire | 2011-2015 | Non-mixture prior | - | Limited subnational data |

| | | | | |
|---|---|---|---|---|
| (CIV) | 2016-2020 | No spatial autoregressive term | - | No subnational data |
| Cameroon (CMR) | 2011-2015 | Standard | - | - |
| | 2016-2020 | Standard | - | - |
| Democratic Republic of the Congo (COD) | 2011-2015 | Standard | - | - |
| | 2016-2020 | Standard | Censoring threshold is 1 | Substantial annual and incremental near-annual observations |
| Republic of the Congo (COG) | 2011-2015 | Standard | - | - |
| | 2016-2020 | Standard | - | - |
| Djibouti (DJI) | 2011-2015 | No spatial autoregressive term | - | No subnational data |
| | 2016-2020 | No spatial autoregressive term | - | No subnational data |
| Eritrea (ERI) | 2011-2015 | No spatial autoregressive term | - | Zero cases |
| | 2016-2020 | No spatial autoregressive term | - | Zero cases |
| Ethiopia (ETH) | 2011-2015 | Standard | - | - |
| | 2016-2020 | Standard | | - |
| Gabon (GAB) | 2011-2015 | No spatial autoregressive term | - | Zero cases |
| | 2016-2020 | No spatial autoregressive term | - | Zero cases |
| Ghana (GHA) | 2011-2015 | Standard | - | - |
| | 2016-2020 | No spatial autoregressive term | - | No subnational data |
| Guinea (GIN) | 2011-2015 | Standard | - | - |
| | 2016-2020 | Standard | - | Limited subnational data |
| Guinea-Bissau (GNB) | 2011-2015 | Non-mixture prior | - | Limited subnational data |
| | 2016-2020 | No spatial autoregressive term | - | Zero cases |
| Equatorial Guinea (GNQ) | 2011-2015 | No spatial autoregressive term | - | Zero cases |
| | 2016-2020 | No spatial autoregressive term | - | Zero cases |
| Kenya (KEN) | 2011-2015 | Standard | - | - |
| | 2016-2020 | Non-mixture prior | - | Improved convergence |
| Liberia (LBR) | 2011-2015 | No spatial autoregressive term | - | No subnational data |
| | 2016-2020 | Standard | - | - |
| Lesotho (LSO) | 2011-2015 | No spatial autoregressive term | - | Zero cases |
| | 2016-2020 | No spatial autoregressive term | - | Zero cases |
| Madagascar (MDG) | 2011-2015 | Non-mixture prior | - | Limited subnational data |
| | 2016-2020 | No spatial autoregressive term | - | Zero cases |
| Mali (MLI) | 2011-2015 | Non-mixture prior | - | Limited subnational data |
| | 2016-2020 | No spatial | - | Zero cases |

| | | | | |
|---|---|---|---|---|
| | | autoregressive term | | |
| Mozambique (MOZ) | 2011-2015 | Non-mixture prior | - | Limited subnational data |
| | 2016-2020 | Standard | - | - |
| Mauritania (MRT) | 2011-2015 | Non-mixture prior | - | Limited subnational data |
| | 2016-2020 | No spatial autoregressive term | - | Zero cases |
| Malawi (MWI) | 2011-2015 | Standard | - | - |
| | 2016-2020 | Standard | - | - |
| Namibia (NAM) | 2011-2015 | Non-mixture prior | - | Limited subnational data |
| | 2016-2020 | No spatial autoregressive term | - | Zero cases |
| Niger (NER) | 2011-2015 | Standard | - | - |
| | 2016-2020 | Standard | - | - |
| Nigeria (NGA) | 2011-2015 | Standard | - | - |
| | 2016-2020 | Standard | Censoring threshold is 1 | Substantial annual and incremental near-annual observations |
| Rwanda (RWA) | 2011-2015 | Standard | - | - |
| | 2016-2020 | No spatial autoregressive term | - | Zero cases |
| Sudan (SDN) | 2011-2015 | No spatial autoregressive term | - | Zero cases |
| | 2016-2020 | Standard | - | - |
| Senegal (SEN) | 2011-2015 | No spatial autoregressive term | - | Limited subnational data |
| | 2016-2020 | No spatial autoregressive term | - | Zero cases |
| Sierra Leone (SLE) | 2011-2015 | Standard | - | - |
| | 2016-2020 | Non-mixture prior | - | Improved convergence |
| Somalia (SOM) | 2011-2015 | Non-mixture prior | - | Improved convergence |
| | 2016-2020 | Standard | - | - |
| South Sudan (SSD) | 2011-2015 | Standard | - | - |
| | 2016-2020 | Standard | - | - |
| Eswatini (SWZ) | 2011-2015 | No spatial autoregressive term | - | Zero cases |
| | 2016-2020 | No spatial autoregressive term | - | No subnational data |
| Chad (TCD) | 2011-2015 | Standard | - | - |
| | 2016-2020 | Standard | - | - |
| Togo (TGO) | 2011-2015 | Non-mixture prior | - | Limited subnational data |
| | 2016-2020 | Non-mixture prior | - | Limited subnational data |
| Tanzania (TZA) | 2011-2015 | Standard | - | - |
| | 2016-2020 | Standard | - | - |
| Uganda (UGA) | 2011-2015 | Standard | - | - |
| | 2016-2020 | Standard | - | - |
| South Africa (ZAF) | 2011-2015 | No spatial autoregressive term | - | Zero cases |

| | | | | |
|---|---|---|---|---|
| | 2016-2020 | No spatial autoregressive term | - | No subnational data |
| Zambia (ZMB) | 2011-2015 | Standard | - | - |
| | 2016-2020 | Standard | - | - |
| Zimbabwe (ZWE) | 2011-2015 | Standard | - | - |
| | 2016-2020 | Standard | - | - |

**Table S4.** Source of unified geographic shapefiles for modeled outputs at the country and second administrative level scales.

| Source | Countries |
|---|---|
| GADM v4.1 (pulled from R package geodata v0.5.9) | Angola, Benin, Botswana, Burkina Faso, Cameroon, Central African Republic, Chad, Côte d'Ivoire, Djibouti, Equatorial Guinea, Eritrea, Eswatini, Gabon, Ghana, Guinea, Guinea-Bissau, Kenya, Lesotho, Liberia, Madagascar, Mali, Mauritania, Mozambique, Namibia, Niger, Nigeria, Republic of the Congo, Rwanda, Senegal, Sierra Leone, Somalia, South Africa, South Sudan, Sudan, Tanzania, Togo, Zambia, Zimbabwe |
| geoboundaries v3.0 (pulled from R package rgeoboundaries v0.0.0.9) | Burundi, Democratic Republic of the Congo, Ethiopia, Malawi, Uganda |

**Table S5.** Public documents from which cholera occurrence data were extracted in the post-2020 period by country, administrative unit level, and time range. Data from these documents were used for the analysis assessing associations between ten-year cholera incidence categories and post-2020 cholera occurrence. While the comprehensive time range represented in the data was from October 2021 to January 2024, the vast majority of data represents cholera occurrence from January 2022 to December 2023.

| Document Name | Country | Admin-level and Time Range |
|---|---|---|
| WHO External Sitrep #1 | Burundi | Admin 1: Jan 2023 - Mar 2023<br>Admin 2: Jan 2023 - Mar 2023 |
| | Democratic Republic of the Congo | Admin 2: Jan 2023 - Mar 2023<br>Admin 3: Jan 2023 - Mar 2023 |
| | Malawi | Admin 1: Mar 2023 - Mar 2023 |
| | Mozambique | Admin 2: Sep 2022 - Mar 2023<br>Admin 3: Sep 2022 - Mar 2023 |
| | South Africa | Admin 2: Jan 2023 - Mar 2023 |
| | Tanzania | Admin 2: Jan 2023 - Mar 2023 |
| | Zambia | Admin 2: Jan 2023 - Mar 2023<br>Admin 3: Jan 2023 - Mar 2023 |
| | Zimbabwe | Admin 1: Jan 2023 - Mar 2023 |
| WHO External Sitrep #2 | Democratic Republic of the Congo | Admin 1: Jan 2023 - Apr 2023<br>Admin 2: Jan 2023 - Apr 2023 |
| | Ethiopia | Admin 2: Jan 2023 - Jan 2023 |
| | Kenya | Admin 1: Jan 2023 - Jan 2023 |
| | Malawi | Admin 1: Apr 2023 - Apr 2023 |

| | | |
|---|---|---|
| | Somalia | Admin 2: Jan 2023 - Jan 2023 |
| | Zimbabwe | Admin 1: Feb 2023 - Apr 2023 |
| WHO External Sitrep #3 | Cameroon | Admin 1: Mar 2023 - May 2023 |
| | Democratic Republic of the Congo | Admin 2: Feb 2023 - Feb 2023 |
| | Ethiopia | Admin 2: Feb 2023 - Feb 2023<br>Admin 3: Feb 2023 - Feb 2023 |
| | Kenya | Admin 1: Feb 2023 - Feb 2023 |
| | Malawi | Admin 1: Apr 2023 - May 2023 |
| | Mozambique | Admin 2: Feb 2023 - May 2023 |
| | Somalia | Admin 2: Feb 2023 - Feb 2023 |
| | South Africa | Admin 2: Feb 2023 - Feb 2023 |
| | Tanzania | Admin 2: Feb 2023 - Feb 2023 |
| | Zambia | Admin 2: Feb 2023 - Feb 2023 |
| | Zimbabwe | Admin 1: Feb 2023 - May 2023 |
| WHO External Sitrep #4 | Burundi | Admin 1: Mar 2023 - Mar 2023<br>Admin 2: Mar 2023 - Mar 2023 |
| | Democratic Republic of the Congo | Admin 2: Mar 2023 - Mar 2023 |
| | Eswatini | Admin 1: Mar 2023 - Mar 2023 |
| | Ethiopia | Admin 2: Mar 2023 - Mar 2023<br>Admin 3: Mar 2023 - Mar 2023 |
| | Kenya | Admin 1: Mar 2023 - Mar 2023 |
| | Mozambique | Admin 2: Mar 2023 - Jun 2023<br>Admin 3: Mar 2023 - Mar 2023 |
| | Malawi | Admin 1: May 2023 - Jun 2023 |
| | Somalia | Admin 2: Mar 2023 - Mar 2023 |
| | South Africa | Admin 2: Feb 2023 - Jun 2023 |
| | Tanzania | Admin 1: Mar 2023 - Mar 2023<br>Admin 2: Mar 2023 - Mar 2023 |
| | Zambia | Admin 2: Mar 2023 - Mar 2023 |
| | Zimbabwe | Admin 1: Mar 2023 - Jun 2023 |
| WHO External Sitrep #5 | Burundi | Admin 2: Apr 2023 - Apr 2023 |
| | Democratic Republic of the Congo | Admin 2: Apr 2023 - Apr 2023 |
| | Eswatini | Admin 1: Apr 2023 - Apr 2023 |
| | Ethiopia | Admin 2: Apr 2023 - Apr 2023<br>Admin 3: Apr 2023 - Apr 2023 |
| | Kenya | Admin 1: Apr 2023 - Apr 2023 |
| | Mozambique | Admin 2: Apr 2023 - Apr 2023<br>Admin 3: Apr 2023 - Apr 2023 |
| | Malawi | Admin 1: Apr 2023 - Jul 2023 |
| | Somalia | Admin 2: Apr 2023 - Apr 2023 |
| | South Africa | Admin 2: Feb 2023 - Jul 2023 |
| | Zambia | Admin 2: Apr 2023 - Apr 2023 |
| | Zimbabwe | Admin 1: Apr 2023 - Apr 2023 |
| WHO External Sitrep #6 | Burundi | Admin 2: May 2023 - May 2023 |
| | Democratic Republic of | Admin 2: May 2023 - May 2023 |

| | | |
|---|---|---|
| | the Congo | |
| | Ethiopia | Admin 2: May 2023 - May 2023<br>Admin 3: May 2023 - May 2023 |
| | Kenya | Admin 1: May 2023 - May 2023 |
| | Mozambique | Admin 1: May 2023 - May 2023<br>Admin 2: May 2023 - Aug 2023 |
| | Malawi | Admin 1: May 2023 - Aug 2023 |
| | Somalia | Admin 2: May 2023 - May 2023 |
| | South Africa | Admin 2: May 2023 - May 2023 |
| | Tanzania | Admin 1: May 2023 - May 2023<br>Admin 2: May 2023 - May 2023 |
| | Zambia | Admin 2: May 2023 - May 2023 |
| | Zimbabwe | Admin 1: May 2023 - May 2023<br>Admin 2: May 2023 - May 2023 |
| WHO External Sitrep #7 | Burundi | Admin 2: Dec 2022 - Sep 2023 |
| | Democratic Republic of the Congo | Admin 2: Jun 2023 - Jun 2023 |
| | Ethiopia | Admin 2: Jun 2023 - Jun 2023<br>Admin 3: Jun 2023 - Jun 2023 |
| | Kenya | Admin 1: Jun 2023 - Jun 2023 |
| | Mozambique | Admin 2: May 2023 - Sep 2023<br>Admin 3: Jun 2023 - Jun 2023 |
| | Malawi | Admin 1: Jun 2023 - Jun 2023 |
| | Somalia | Admin 2: Jun 2023 - Jun 2023<br>Admin 3: Jun 2023 - Jun 2023 |
| | South Africa | Admin 1: Jun 2023 - Jun 2023 |
| | Zambia | Admin 2: Jun 2023 - Jun 2023 |
| | Zimbabwe | Admin 1: Jun 2023 - Jun 2023 |
| WHO External Sitrep #8 | Burundi | Admin 2: Jul 2023 - Jul 2023 |
| | Democratic Republic of the Congo | Admin 2: Jul 2023 - Jul 2023 |
| | Ethiopia | Admin 2: Jul 2023 - Jul 2023<br>Admin 3: Jul 2023 - Jul 2023 |
| | Kenya | Admin 1: Jul 2023 - Jul 2023<br>Admin 3: Jul 2023 - Jul 2023 |
| | Mozambique | Admin 2: Jul 2023 - Oct 2023<br>Admin 3: Jul 2023 - Oct 2023 |
| | Malawi | Admin 1: Jul 2023 - Jul 2023 |
| | Somalia | Admin 2: Jul 2023 - Jul 2023<br>Admin 3: Jul 2023 - Jul 2023 |
| | South Africa | Admin 2: Jul 2023 - Jul 2023<br>Admin 3: Jul 2023 - Jul 2023 |
| | Tanzania | Admin 1: Jul 2023 - Jul 2023<br>Admin 2: Jul 2023 - Jul 2023 |
| | Uganda | Admin 1: Jul 2023 - Jul 2023 |
| | Zimbabwe | Admin 1: Jul 2023 - Oct 2023 |
| WHO External Sitrep #9 | Burundi | Admin 2: Aug 2023 - Aug 2023 |
| | Democratic Republic of the Congo | Admin 2: Aug 2023 - Aug 2023 |
| | Ethiopia | Admin 1: Aug 2023 - Aug 2023 |

| | | |
|---|---|---|
| | Kenya | Admin 1: Aug 2023 - Aug 2023 |
| | Mozambique | Admin 2: Aug 2023 - Nov 2023<br>Admin 3: Aug 2023 - Nov 2023 |
| | Malawi | Admin 1: Aug 2023 - Aug 2023 |
| | Somalia | Admin 2: Aug 2023 - Aug 2023<br>Admin 3: Aug 2023 - Aug 2023 |
| | Sudan | Admin 1: Aug 2023 - Aug 2023 |
| | Uganda | Admin 1: Aug 2023 - Aug 2023<br>Admin 2: Aug 2023 - Aug 2023 |
| | Zambia | Admin 2: Aug 2023 - Aug 2023 |
| | Zimbabwe | Admin 1: Aug 2023 - Nov 2023 |
| WHO External Sitrep #10 | Burundi | Admin 1: Sep 2023 - Sep 2023<br>Admin 2: Sep 2023 - Dec 2023 |
| | Democratic Republic of the Congo | Admin 2: Sep 2023 - Jan 2024<br>Admin 3: Oct 2023 - Dec 2023 |
| | Ethiopia | Admin 1: Sep 2023 - Dec 2023 |
| | Kenya | Admin 1: Sep 2023 - Nov 2023 |
| | Mozambique | Admin 2: Sep 2023 - Dec 2023<br>Admin 3: Sep 2023 - Dec 2023 |
| | Malawi | Admin 1: Sep 2023 - Dec 2023 |
| | Somalia | Admin 2: Sep 2023 - Dec 2023 |
| | Sudan | Admin 1: Sep 2023 - Dec 2023 |
| | Tanzania | Admin 1: Oct 2023 - Dec 2023 |
| | Zambia | Admin 2: Sep 2023 - Dec 2023 |
| | Zimbabwe | Admin 1: Sep 2023 - Dec 2023 |
| AFRO Cholera Bulletin.49 | Cameroon | Admin 1: Oct 2021 - Jan 2024 |
| | Togo | Admin 1: Dec 2023 - Dec 2023 |
| | Zambia | Admin 1: Jan 2024 - Jan 2024 |
| An update of Cholera outbreak in Nigeria_221222_52 | Nigeria | Admin 1: Jan 2022 - Dec 2022 |
| An update of Cholera outbreak in Nigeria_221223_52 | Nigeria | Admin 1: Jan 2023 - Dec 2023 |
| Weekly Bulletin on Outbreaks and Other Emergencies - WHO African Region | Tanzania | Admin 2: Sep 2023 - Oct 2023 |
| South Sudan Cholera Situation Report_Issue #39 | South Sudan | Admin 2: Feb 2023 - Apr 2023 |
| WHO Sudan Outbreaks dashboard | Sudan | Admin 3: Jun 2023 - Jan 2024 |

**Table S6.** Summary of full-year observations by period. This summary represents all model input observations, including imputed national-level observations. Observations with a time fraction greater than or equal to eight months (0.65 years) were considered full-year observations. Observations with shorter time fractions were considered as right-censored in the model likelihood.

| Period | Full-year observations (%) | Total observations |
|---|---|---|
| 2011-2015 | 13,324 (89) | 14,940 |
| 2016-2020 | 8,756 (57) | 15,271 |

| | | |
|---|---|---|
| 2011-2020 | 22,080 (73) | 30,211 |

### Supplementary Material

#### Cholera data sources and data processing

Cholera data were extracted from a database and processed before being passed to a statistical mapping model to produce gridded estimates of mean annual incidence (Figure S1). The main data processing steps consist of temporal aggregation to the yearly time scale, identifying temporally censored observations (those spanning less than 8 months of a year), filtering observations that do not contribute to the likelihood, imputation of limited national-level observations, and assigning observation-linked geographic areas ("locations") to the spatial modeling grid. After modeling, the resulting gridded estimates undergo post-processing to produce estimates for unified, non-overlapping administrative units.

All countries in Africa that had at least one national-level report of suspected cholera (including zero) in both periods of analysis were modeled (Table S2). Following this criterion, 11 of 54 countries in Africa were excluded (Algeria, Cape Verde, Comoros, Egypt, Gambia, Libya, Mauritius, Morocco, Sao Tome and Principe, Seychelles, Tunisia).

*Cholera data collection and data template*

All cholera surveillance and alert documents were systematically scraped for all reported counts of suspected cholera (henceforth "observations") that were explicitly linked to date ranges and geographical areas (henceforth "locations") and were thought to represent all cases reported in a specific space-time unit (e.g., not representing just a subset of cases, such as age- or sex-stratified counts). Location names were systematically verified and associated geographic shapefiles were identified with a standard location audit protocol, which consisted of searches on reputed websites and resources and comparison to locations that already existed in the Cholera Taxonomy database [1,2]. Metadata, source documents, shapefiles, and observations were then added to the global cholera surveillance database. Each observation contained the following information: location shapefile, date range, number of suspected cases, and time fraction (tfrac) within a calendar year, which is calculated from the date range.

*Temporal aggregation*

Our statistical mapping model aimed to infer mean annual incidence rates, so we sought to aggregate observations to the annual time resolution. As observations may exist for arbitrary locations and date ranges, non-overlapping observations that were consecutive in time were aggregated if they were in the same location, calendar year, and source document. If a location had multiple observations of suspected cholera for the same time bounds within a same data source, we included the observation with the largest case count in the aggregated observation; the implicit assumption here is that cases are more likely to be underreported than overreported so we give preference to higher case count reports. This resulted in a set of aggregated observations per location, year, and data source, and the corresponding fraction of calendar year they covered, that were used as model inputs.

*Identifying time-censored observations*

Our modeling framework did not assume that cholera incidence was homogenous throughout the year (cf. *Inference modeling framework*), and we therefore differentiated full-year observations from partial-year observations in the model likelihood. Partial-year observations were considered to be right-censored if they spanned less than eight months (0.65 years). We dropped all right-censored observations with zero suspected cases, as these have a likelihood of 1 and therefore do not contribute to the model likelihood.

*Observation filtering*

Observations were dropped from inclusion in the model if they have a likelihood of 1 (and therefore would not contribute to the likelihood) or were otherwise not informative to the model. While some of these decisions are discussed elsewhere in the SM, broadly speaking, observations that were removed include those that: 1) were not associated with a geographic shapefile, 2) were ADM0 (country-level) observations that spanned multiple years, 3) exactly duplicated observations prior to temporal aggregation, 4) had the exact same location and time bounds but reported fewer cases than another temporally aggregated observation from the same data source, 5) time-censored observations that reported zero cases, 6) time-censored observations at the ADM0 level that had less than half the reported cases as full-year ADM0 observations in that timeslice, or 7) had zero population according to the WorldPop raster.

*National-level data imputation*

At least one country-level annual observation was sought for every country-year combination modeled. This imposed a critical constraint to bound modeled incidence rate estimates, particularly when fitting a model to data with only censored observations or only subnational observations and incomplete spatial coverage across the country.

When a country-level observation was not found in a given year, imputation of a country-level annual observation was performed. If there were no observations at any spatial scale available for that year, a zero-case observation was imputed, thus assuming that absence of data in a year corresponded to a report of 0 cases for that country. If subnational observations were available and they covered a non-overlapping spatial area that represents at least 10% of the country population, a mean tfrac-adjusted incidence rate was computed across all subnational observations and multiplied by the country population to impute a country annual observation. If subnational observations were available and they covered a spatial area representing less than 10% of the country population, an observation was imputed as the maximum of the sum of cases across all unique data source and administrative unit level combinations. If only censored national observations and no subnational observations were available, the maximum value censored observation was imputed.

In the end, a limited number of observations were imputed (109 imputed relative to 30,102 non-imputed observations) in order to ensure that all modeled countries had at least one country-level observation per year, a data feature that was found to improve model stability and performance. Of these, zero-case observations were added when no annual country-level report was found (96 imputed observations in 21 countries). When subnational or censored national annual reports were available, non-zero-case observations were aggregated to impute an annual country-level report (13 imputed observations in 10 countries).

*Geographic linkage of observations to modeling grid*

Our statistical modeling framework was applied to a space-time modeling grid (Figure S2), where the space dimension was composed of 20 km by 20 km grid cells that overlapped with a given country's geographic shapefile and had a population greater than zero according to the associated WorldPop gridded population estimates in that year; the time dimension was represented in annual time slices.

Observations of suspected cholera were associated with space-time cells in the modeling grid according to their geographic shapefiles and date ranges. In the space dimension, observations were geographically linked to all 20 km by 20 km grid cells that intersected the observation's geographic shapefile. When grid cells were only partially covered by the observation's geographic shapefile (eg., grid cells at country borders), we computed and assigned a spatial fraction (sfrac) to the grid cell-shapefile pairs. The sfrac value was calculated as the sum of the 1 km by 1 km grid cell population (after aligning the 1 km by 1 km WorldPop gridded population estimates to the 20 km by 20 km spatial modeling grid) that intersected the observation's geographic shapefile divided by the total 20 km by 20 km grid cell population. In the time dimension, observations were mapped to all annual time slices that overlapped with the observation date range.

We removed cell-shapefile linkages with small overlaps in order to improve the smoothness of model estimates at shapefile borders. Spatial grid cells that overlapped with an observation's geographic shapefile with a population-weighted spatial fraction below 0.05 were removed from being associated with the shapefile. To improve the smoothness of model estimates at country borders, we removed spatial grid cells from the space-time modeling grid if the grid-cell sfrac was less than 0.3.

#### Incidence modeling framework

The goal of our modeling framework was to produce gridded estimates of mean annual incidence of suspected cholera by harnessing potentially overlapping data from heterogeneous spatial and temporal resolutions and multiple sources. In particular, the framework accounted for misalignment in spatial and temporal resolutions both across data sources and between observed case counts and intended outputs. This model expanded on a previously published approach [1]. We first describe the base statistical model and add complexity that improves the base model's ability to handle challenges presented by the real-world observation data. At the end, we present the full final "standard" model structure and deviations from this standard model structure, which were deployed in country-periods described in Table S3.

*Base model*

To estimate mean annual incidence across a period of $T$ years ($T$ annual time slices), we first must model annual cholera incidence estimates corresponding to a "modeling time resolution" of 1 year. In a simple scenario, suppose that all observations have a duration of 1 year, which means that the “observation time resolution” always equals the modeling time resolution. To model space-time incidence rates over a spatial domain that covers the area of interest across the *T* years, we defined a modeling space-time grid with a time resolution of 1 year for a given gridded spatial resolution, i.e., each space-time grid-cell (s,t) spans one year t and a spatial grid cell s.

Observation-level cases can then be modeled as:

$$c_i = \sum_{S_{i,s,t}} \lambda_{s,t} \phi_{i,s} pop_{s,t} ,$$

$$log(\lambda_{s,t}) = \gamma + \omega_s + \eta_t,$$

where $c_i$ represents the modeled mean number of cases for observation *i*, $S_{i,s,t}$ is the set of space-time grid cells intersecting observation *i*, $\lambda_{s,t}$ is the annual incidence rate in space-time grid cell $s,t$, $\phi_{i,s}$ is the population-weighted spatial fraction of grid cell *s,t* that is covered by the observation location, and $pop_{s,t}$ is the total population in grid cell *s,t*. Grid cell incidence rates were modeled with a log link as the sum of the offset $\gamma$, which is the expected incidence rate across the space-time modeling grid, spatial random effect $\omega_s$, and yearly random effect $\eta_t$.

The expected incidence rate $\gamma$ was calculated as the population-weighted mean of the implied incidence rate (time-adjusted reported cases of full-year observations divided by location population) across all full-year observations contributing to the model (cf. *Identifying time-censored observations*). If the expected incidence rate was less than 0.01 per 100,000 population, it was changed to be 1E-7.

Observation $y_i$ is then linked to modeled cases through an observation model. For instance, in the simplest setting, one can assume that observations follow a Poisson distribution:

$$y_i \sim Poisson(c_i).$$

However, a Poisson model does not reflect the heterogeneity in case-counts observed in the data, thereby necessitating a more elaborate observation process model. We expand on the observation process below.

*Prior on the spatial random effect ($\omega_s$)*

To capture spatial variability in the incidence rates and produce spatially smooth maps, we introduced spatial random effects into the model at the grid-cell level. We assumed that the spatial random effect $\omega_s$ was constant across the *T* time slices to reduce the number of parameters that must be estimated from a model that may have limited observations in any given time slice. We acknowledge that this model may not adequately capture situations where the spatial autocorrelation in cholera cases changes across time slices. In such scenarios, the estimates of $\omega_s$ will represent the average spatial variability across all the time slices.

In our model, the joint distribution of $\omega_s$ for all spatial grid cells $s$ is specified as a directed acyclic graph auto-regressive (DAGAR) prior. The DAGAR model was demonstrated to have improved model performance, interpretability of parameters, and computational efficiency over other spatially-smooth priors that are traditionally used in disease mapping (e.g., conditional auto-regressive (CAR) prior) [3]. The DAGAR prior can be specified via a sequence of simple conditional normal distributions. Specifically, the conditional distribution of $\omega_s$, conditional on its directed neighbors on the grid, follows a normal distribution with mean $\mu_{\omega_s}$ and standard deviation $\sigma_{\omega_s}$:

$$\omega_s \sim Normal(\mu_{\omega_s}, \sigma_{\omega_s}),$$

$$\mu_{\omega_s} = \frac{\rho}{(1+(nn_s-1)\rho^2)} \sum_{u \in \Omega_s} \omega_u,$$

$$\sigma_{\omega_s} = \xi_{\sigma_w} \sqrt{\frac{(1-\rho^2)}{(1+(nn_s-1)\rho^2)}},$$

where $\rho$ is the strength of the spatial autocorrelation between grid cells, $nn_s$ is the number of neighbors of cell $s$, and $\Omega_s$ is the set of neighbors to cell $s$. We denote the DAGAR prior as $\omega \sim DAGAR(\rho, \xi_{\sigma_w})$ where $\omega$ is a vector containing $\omega_s$ for all the spatial grid cells $s$.

*Prior on the temporal random effect ($\eta_t$)*

While the yearly temporal random effects were initially assumed to be independent, we imposed a zero-sum constraint to improve identifiability of these parameters and enforced a marginal standard normal prior on the set of these terms [4]. Briefly, the approach applies a QR decomposition on the covariance matrix of the yearly random effect to obtain a set of random variables with a zero-sum constraint and marginal standard deviations of 1. In practice, priors are set on *T - 1* independent random effects, and the random effect of the *T*th time slice is computed from them.

*Expansion for partial-year observations*

Partial-year observations (i.e., those with tfrac < 0.65 within a given annual modeling time slice) were treated as right-censored in the likelihood (cf. *Identifying time-censored observations*). Because we assumed that incidence rates were non-homogeneous within a given annual time slice, we chose to treat partial-year observations as right-censored observations of the annual counts as opposed to performing an extrapolation to represent a full year. In other words, we make no assumptions beyond that the number of cases in the full-year modeling time slice would be at least as large as the number of observed cases in the partial-year observation. The observation model likelihood for partial-year observations was:

$$L(y_i) = Pr(Y \geq y_i | c_i),$$

so in the case of the Poisson observation model, the likelihood is:

$$L(y_i) = 1 - CDF_{Poisson}(y_i | c_i).$$

*Expansion for overdispersed observation data*

Examination of the observation data determined that a Poisson observation model would not be sufficient to account for the overdispersion observed in many country-period models and across different administrative unit levels. Consequently, we accounted for overdispersion with a negative binomial observation likelihood:

$$y_i \sim NegBinom(c_i | \tau_{A[i]}),$$

where $\tau$ is the overdispersion parameter that defines the relationship of the mean $c$ to the variance:

$$variance \ = c + c^2/\tau.$$

To account for expected differences in overdispersion across administrative level reporting, the model allowed for different overdispersion parameters by observation administrative unit level *A[i]*. The overdispersion parameter ($\tau_{A0}$) was fixed at the country level (A0), but inferred for all other administrative unit levels.

*Complete standard model formulation*

The final standard model followed a hierarchical structure, such that the process model was defined:

$$c_i = \sum_{S_{i,s,t}} \lambda_{s,t} \phi_{i,s} pop_{s,t} \ ,$$

$$log(\lambda_{s,t}) = \gamma + \omega_s + \eta_t,$$

and the observation model was defined for full-year and partial-year observations:

$$Pr(y_i | c_i) \ = NegBinom(c_i | \tau_{A[i]}), \qquad if\ \Phi_{i,t} \geq a$$

$$Pr(y_i | c_i) = 1 - CDF_{NegBinom}(y_i | c_i, \tau_{A[i]}) \qquad if\ \Phi_{i,t} < a,$$

where $a \in [0.65, 1]$ is range of time fractions $\Phi_{i,t}$ for which an observation *i* is considered to represent the full year *t*.

We used the following hyperpriors for the spatial random effects:

$$\omega \sim DAGAR(\rho, \xi_{\sigma_w}),$$

$$\rho \sim Beta(5, 1.5),$$

$$Pr(\xi_{\sigma_w}) = \theta f_{Normal}(\xi_{\sigma_w} | 10, \sigma'_{\omega,1}) + (1 - \theta) f_{Normal}(\xi_{\sigma_w} | 0, \sigma'_{\omega,2}),$$

$$\theta \sim Beta(1, 3),$$

$$\sigma'_{\omega,1} \sim Half\ normal(0, 2),$$

$$\sigma'_{\omega,2} \sim Half\ normal(0, 0.5),$$

where $Pr(\xi_{\sigma_w})$ represents a mixture prior on the standard deviation scaling constant ($\xi_{\sigma_w}$), which is the sum of two normal distribution densities ($f_{Normal}$), one centered at 0 and the other centered at 10, weighted by the mixture parameter ($\theta$). The hyperpriors on the standard deviation of the normal distribution densities contributing to the mixture prior are represented by $\sigma'_{\omega,1}$ and $\sigma'_{\omega,2}$. This mixture prior reflects the possibility that different models may have high or low magnitude of spatial variability.

We used the following prior for temporal random effects:

$$\eta'_{[1:T-1]} \sim Normal(0, \frac{1}{\sqrt{1-1/T}}),$$

where $\eta'_{[1:T-1]}$ is multiplied by the QR matrix (cf. *Prior on the temporal random effects*) to yield the $\eta_{[1:T]}$ yearly random effects for all *T* time slices whose sum is enforced to be 1.

For the observation model, the overdispersion term for administrative unit level 0 (*A0*) observations was fixed at 100 or 1000, which corresponded to a moderate amount of overdispersion for case counts of that magnitude. We used the following priors for the overdispersion term in the observation model:

$$\tau_{A0} = 100 \quad if\ max(y_{i,A0}) \leq 5000\ ,$$

$$\tau_{A0} = 1000 \quad if\ max(y_{i,A0})\ >\ 5000\ ,$$

$$\frac{1}{\tau_{A>0}} \sim Half\ normal(0, 1),$$

where *A>0* refers to administrative level units below the national level.

*Model formulation without spatial autoregressive term*

For country-periods with no subnational observations or only zero-case observations, we removed the spatial random effect from the process model. For this model deviation, spatial random effect priors were removed and the process model was as follows:

$$c_i = \sum_{S_{i,s,t}} \lambda_{s,t} \phi_{i,s} pop_{s,t}\ ,$$

$$log(\lambda_{s,t})\ =\ \gamma\ +\ \eta_t.$$

*Model selection and model formulation with non-mixture prior*

All country-periods with at least one subnational non-zero-case observation were first attempted with the standard model formulation. We found that models with limited subnational data had poor model fit and convergence due to identifiability issues in the spatial autocorrelation strength parameter $\rho$ in the spatial random effect $\omega$. The standard model employed a mixture prior on the scaling factor ($\xi_{\sigma_w}$) of the standard deviation of $\omega$ ($\sigma_{\omega_s}$) to account for possible bimodality in the strength of spatial autocorrelation (i.e., country-periods may have high or low $\rho$ depending on the data).

For country-periods with limited subnational data, model convergence improved when the mixture prior on $\xi_{\sigma_w}$ was replaced with a unimodal prior that favors a higher $\sigma_{\omega_s}$ and therefore lower $\rho$. The scaling constant priors for this model deviation reduced to:

$$\xi_{\sigma_w} \sim Half\ normal(5, 0.5)$$

*Incidence modeling post-processing*

Our modeling framework produced 20 km by 20 km gridded mean annual incidence estimates for each country and time period (1000 posterior samples). Grid cell estimates were then aggregated to administrative unit level 0 (country or ADM0) and administrative unit level 2 (district or ADM2) scales according to standardized sets of non-overlapping country-unified shapefiles. These were obtained for each modeled country from GADM [25] or geoBoundaries [24] after an additional quality assessment (Table S4). Lesotho did not have ADM2 shapefiles from a standard source, so these were processed to the administrative unit level 1 scale instead.

A set of continent- and region-wide posterior samples were assembled by summing across a single random posterior predictive sample of each country model; mean and 95% credible intervals (CrIs) were then calculated from the summed results, thus yielding 1000 continent-wide and region-wide posterior predictive samples that are internally consistent.

Multiple outputs were calculated at continent, regional, ADM0, and ADM2 scales for each model in post-processing: mean annual incidence (cases per year), mean annual incidence rate (cases per population per year), incidence rate ratio (abbreviated "IRR", calculated as mean annual incidence rate in 2016-2020 divided by that in 2011-2015), assignment of ADM2 units to 5-year and 10-year incidence categories, and population in ADM2 units in 5-year and 10-year incidence categories. Mean annual incidence and mean annual incidence rate posterior mean and 95% CrI estimates were calculated across 1000 posterior predictive samples at the relevant spatial scale. To estimate IRRs, the mean and 95% CrIs were calculated across all pairwise ratios of 1000 posterior predictive samples in each period (i.e., evaluated across the distribution of $1000^2$ samples – all 2016-2020 samples were pairwise-divided by all 2011-2015 samples). IRRs were deemed statistically significant if the 95% CrIs did not cross one.

Number of people living in ADM2 units by 5-year incidence categories at continent and region scales were estimated across the summed 1000 continent- and region-wide posterior samples described above (corresponds to results in Fig 3A and Figure S17). The mean and 95% CrIs for each 5-year incidence category were calculated across the 1000 posterior samples. Consequently, the variability in these estimates reflects variation in ADM2 incidence category assignment across samples.

Assignment of ADM2 units to specific 5-year incidence categories was performed in a two-step procedure (corresponds to results in Fig 3B and Figure S18). First we determined incidence categories for each 20 km by 20 km modeling grid cell and posterior sample, and retained the highest incidence category encompassing at least 10% of the unit's population or 100,000 people in 2020. Then, ADM2 units were assigned to the lowest incidence category with at least 50% posterior cumulative probability of assignment at or above that level (e.g., ADM2 unit was assigned to the 50-100 cases per 100,000 category if ≥ 50% of posterior samples categorized the ADM2 unit in the 50-100 or ≥100 cases per 100,000 people categories). Thus, the assignment of an ADM2 unit to an incidence category already factors in the variability in assignment across samples. We note that summing the number of people in all ADM2 units by 5-year incidence category would not yield the same number as the continent-wide mean estimate of population living in the ADM2 category.

Assignment of ADM2 units to 10-year incidence categories was based directly on their 2011-2015 and 2016-2020 incidence category assignments. We defined four 10-year incidence categories: "sustained high" for ADM2 units classified as high incidence (≥10 cases per 100,000 people per year) in both periods, "history of high" for ADM2 units classified as high incidence in at least one period, "sustained low" for ADM2 units classified as <1 case per 100,000 per year in both periods, and "history of moderate" for all other combinations.

#### Analysis of 2022-2023 cholera outbreak occurrence

A second extracted dataset and modeling framework were used to estimate the association between 2011-2020 incidence categories and the probability of reporting suspected cholera occurrence in the 2022-2023 period.

##### *Extraction and spatial linkage of cholera occurrence data*

Fourteen situation report documents (Table S5) with map images of cholera occurrence in the post-2020 period were loaded into the QGIS geographic information system software (version 3.28.12) and overlaid with the standardized set of country-unified shapefiles (Table S4). Each image was georeferenced to the country-unified shapefiles as a basemap with country borders as control points. After aligning the administrative unit boundaries, we manually added centroids to extract point locations for each administrative unit and added attributes to identify the administrative unit level, confidence about the certainty of the administrative unit level, presence of reported cholera cases, and the time range represented by the map.

We spatially joined extracted locations with cholera occurrence to the set of unique ADM2 units used to summarize the cholera incidence mapping results. Cholera occurrence was extrapolated to ADM2 units if cholera was reported in ADM3 scale units or below.

##### *Base statistical model*

This analysis aimed to estimate the association between 10-year incidence categories and the probability of reporting suspected cholera occurrence in the 2022-2023 period. For all locations that were modeled, those that reported cholera were indexed with *j* and those that did not report cholera were indexed with $k$ .

For ADM2 locations that reported cholera occurrence, the likelihood is:

$$L(y_{j,A2} = 1) = p_{j,A2}\phi_{j,A2} \text{ and}$$

$$logit(p_{j,A2}) = \alpha + \beta_{j,A2} ,$$

where $y_{j,A2}$ is the reported cholera occurrence status extracted from the situation report documents in location *j* which is at the ADM2 level ($A2$), $p_{j,A2}$ is the probability of true cholera occurrence, $\phi_{j,A2}$ is the probability of reporting cholera if it is present (sensitivity of cholera detection), $\alpha$ is the model intercept, and $\beta_{j,A2}$ is the effect of the 10-year incidence category in the ADM2 unit. Importantly, the model assumes that all reported cholera is a true instance of cholera occurrence (i.e., no false positives).

As the absence of reported occurrence may be due to lack of cholera occurrence or lack of reporting, we treated the absence of reported occurrence as missing data and marginalized out all possible reporting statuses to estimate the underlying true cholera occurrence status. For ADM2 locations that did not report cholera, the likelihood reads:

$$L(y_{k,A2} = 0) = p_{k,A2}(1 - \phi_{k,A2}) + (1 - p_{k,A2}) = 1 - p_{k,A2}\phi_{k,A2} \text{ and}$$

$$logit(p_{k,A2}) = \alpha + \beta_{k,A2} ,$$

where $p_{k,A2}$ is the probability of true cholera occurrence in ADM2 unit $k$, $(1 - \phi_{k,A2})$ is the probability of not reporting cholera if it is indeed present, and $\beta_{k,A2}$ is the effect of the 10-year incidence category in the ADM2 unit.

Reports of cholera occurrence in ADM2 locations could therefore be modeled with a Bernoulli distribution:

$$y_{i,A2} \sim Bernoulli(p_{i,A2}\,\phi_{i,A2}),$$

where *i* represents any location regardless of cholera reporting status.

*Adding higher administrative unit level observations*

As some occurrence data was only available at the ADM1 or ADM0 (country) level, these observations were integrated into the model:

$$y_{i,A<2} \sim Bernoulli(\eta_{i,A<2}) \text{ and}$$

$$\eta_{i,A<2} = 1 - \prod_{i,A2 \in S,A2} 1 - p_{i,A2}\phi_{i,A2},$$

where *S,A2* represents the set of *i,A2* ADM2 units contained within the location *i,A<2* which is above the ADM2 level, and $\eta_{i,A<2}$ is the probability of reported cholera occurrence in the higher administrative unit level location *i,A<2* .

*Hierarchical country and region-level priors*

We assumed that the association between 10-year incidence categories and the probability of cholera occurrence may vary across countries and regions (e.g., Eastern Africa). We accounted for these geographic differences by setting hierarchical priors, such that priors for the association of the 10-year incidence category and probability of true cholera occurrence in location $i$, which is contained within country $c$ and region $r$ were defined as:

$$\beta_c^m \sim Normal(\mu_{\beta,r}^m, \sigma_{\beta,r}^m) \text{ and}$$

$$\mu_{\beta,r}^m \sim Normal(\mu_{\beta}^m, \sigma_{\beta}^m),$$

where $m$ denotes the 10-year incidence category associated with location $i$, $\mu_{\beta,r}^m$ and $\sigma_{\beta,r}^m$ are regional-level mean and standard deviation of the 10-year incidence category effect β, and $\mu_{\beta}^m$ and $\sigma_{\beta}^m$ are hyperpriors for the mean and standard deviation of the 10-year incidence category effect.

Hierarchical priors were also assumed for the cholera detection sensitivity parameters $\phi$, which had analogous relationships on a logit scale:

$$logit(\phi_c) \sim Normal(\mu_{logit(\phi),r}, \sigma_{logit(\phi),r}) \text{ and}$$

$$\mu_{logit(\phi),r} \sim Normal(\mu_{logit(\phi)}, \sigma_{logit(\phi)}).$$

*Model priors and hyperpriors*

We used the following priors:

$$\sigma^{m}_{\beta,r} \sim Half\ normal(0, 2.5),$$
$$\mu^{m}_{\beta} \sim Normal(0, 2),$$
$$\sigma^{m}_{\beta} \sim Half\ normal(0, 2.5),$$
$$\sigma_{logit(\phi),r} \sim Half\ normal(0, 1),$$
$$\mu_{logit(\phi)} \sim Normal(1.5, 5),$$
$$\sigma_{logit(\phi)} \sim Half\ normal(0, 1).$$

##### Assessing intervention reach when prioritizing targets by cholera incidence

We assessed the potential reach of interventions when targeted based on cholera incidence through two analysis types. Both analyses ranked ADM2 units by decreasing incidence category and decreasing population size within incidence categories as a simplification of how intervention targets might be prioritized using cholera incidence data. “Prospective” targeting used past incidence categories to target future interventions, while “oracle” targeting prioritized interventions based on incidence categories from the same period.
In the first analysis, we assessed the proportion of mean annual 2016-2020 cholera cases that would have been reached by interventions had 2011-2015 (“prospective”) or 2016-2020 (“oracle”) incidence categories been used for targeting. We also assessed the proportion of mean annual 2011-2015 cholera cases that would have been reached by interventions had 2011-2015 incidence categories been used for targeting (“oracle” only).

In the second analysis, we examined the proportion of population living in ADM2 units with modeled 2022-2023 cholera occurrence (modeled according to the above described statistical analysis) that would have been reached by interventions had 2011-2015, 2016-2020, 2011-2020 incidence categories (“prospective”) been used for targeting. These three strategies were compared to an “oracle” targeting strategy where ADM2 units with 2022-2023 cholera occurrence were ranked in decreasing order of population size.

---

2 Johns Hopkins Cholera Dynamics Team. Cholera Taxonomy.

https://cholera-taxonomy.middle-distance.com/ (accessed June 12, 2024).

3 Datta A, Banerjee S, Hodges JS, Gao L. Spatial disease mapping using directed acyclic graph auto-regressive (DAGAR) models. *Bayesian Anal* 2019; **14**: 1221–44.

4 Goodman A. Test: Soft vs Hard sum-to-zero constrain + choosing the right prior for soft constrain. The Stan Forums. 2018; published online Sept 24. https://discourse.mc-stan.org/t/test-soft-vs-hard-sum-to-zero-constrain-choosing-the-right-prior-for-soft-constrain/3884/31 (accessed May 2022, 2023).
